# Estimating epidemic dynamics with genomic and time series data

**DOI:** 10.1101/2023.08.03.23293620

**Authors:** Alexander E. Zarebski, Antoine Zwaans, Bernardo Gutierrez, Louis du Plessis, Oliver G. Pybus

## Abstract

Accurately estimating the prevalence and transmissibility of an infectious disease is an important task in genetic infectious disease epidemiology. However, generating accurate estimates of these quantities, that are informed by both epidemic time series and pathogen genome sequence data, is a challenging problem. While birth-death processes and coalescent-based models are popular methods for modelling the transmission of infectious diseases, they both struggle (for different reasons) with estimating the prevalence of infection.

Here we extended our approximate likelihood, which combines phylogenetic information from sampled pathogen genomes and epidemiological information from a time series of case counts, to estimate historical prevalence (in addition to the effective reproduction number). We implement this new method in a BEAST2 package called Timtam. In a simulation study our approximation is well-calibrated and can recover the parameters of simulated data.

To demonstrate how Timtam can be applied to real data sets we carried out empirical analyses of data from two infectious disease outbreaks: the outbreak of SARS-CoV-2 onboard the Diamond Princess cruise ship in early 2020 and poliomyelitis in Tajikistan in 2010. In both cases we recover estimates consistent with previous analyses.

## Introduction

In the field of genetic infectious disease epidemiology, there are two key common questions: “how many people are infected?” (i.e. what is the prevalence?) and “how transmissible is this pathogen?” (i.e. what is its effective reproduction number?) Prevalence of infection is the number of individuals infected at a given time and the effective reproduction number is defined as the average number of secondary infections per infectious individual at a given time.

Birth-death processes are an increasingly popular family of models for describing the transmission of infectious diseases, in part because they capture the mechanism of the process and are amenable to analysis. In the birth-death process, *births* represent new infections and *deaths* the end of an infectious period. In its Bayesian phylogenetic implementation, the birth-death process enters the analysis as a prior model for the reconstructed phylogeny (the so-called *tree prior*.) Kendall, 1948 first demonstrated how to use generating functions to describe birth-death processes when modelling infectious disease. Later, Nee et al., 1994 connected that process to the number of observed species in a phylogeny, and Stadler, 2010 and Stadler et al., 2012 demonstrated how this idea can be applied to the analysis of phylogenies of pathogen genomes.

Coalescent models offer a computationally and mathematically convenient alternative to birth-death models and are often used to analyse viral genomes (Pybus et al., 2000; Volz et al., 2013). However, a number of assumptions must be made in order to apply coalescent models to epidemiological problems and these assumptions may not always be met. Further, while there are exceptions, e.g. Parag et al., 2020 and Volz, 2012, coalescent models typically lack an explicit sampling model, and will estimate an effective population size, *N*_*e*_, rather than the true population size. These aspects can complicate matters in an epidemiological setting.

As with coalescent models, it is difficult to estimate the true population size with birth-death models, and it is usually not undertaken (again, there are exceptions, e.g. Kühnert et al., 2014). This difficulty stems, in part, from the implicit assumption of an infinite susceptible population in most birth-death models. Two remedies offered are the use of parametric models with a finite population, and nonparametric models with time-varying rate parameters. Parametric models impose additional structure, which may require strong simplifying assumptions and can make analysis computationally intractable. Nonparametric models achieve additional flexibility by representing parameters — particularly the birth rate — as piece-wise constant functions (Stadler et al., 2013). In an epidemiological context, to adjust for the depletion of the susceptible pool, the model allows the birth rate to decline over time. While convenient, nonparametric estimates make it difficult to distinguish if a decline in apparent birth rate is due to depletion of the susceptible pool or changes in behaviour, e.g. due to non-pharmaceutical interventions.

Incorporating unsequenced case data into phylodynamic analyses in a principled way is a long-standing challenge for the field (Vaughan et al., 2019). Typically, only a small number of cases are sequenced. For example, no country with a sizeable COVID-19 outbreak sequenced *>* 20% of reported cases and most sequenced *<* 5%. Among low and middle income countries this number is often *<* 1%. Note that these numbers are for *reported* cases and not the true number of infections (Brito et al., 2022). Purely phylodynamic methods rely on this small subset of sequenced cases to estimate epidemiological dynamics, taking the stance that genomic sequences contain useful information and can facilitate reconstructing the dynamics of transmission, even before the first case in an outbreak was identified. Nevertheless, the vast amount of unsequenced case data is also informative and can help to refine estimates of epidemic parameters (Rasmussen et al., 2011; Judge et al., 2023). The calculations needed to simultaneously analyse both sources of data in an integrated framework are well-known (see for example Manceau et al., 2020), however existing ways to compute them are computationally intractable for all but the smallest outbreaks (Andréoletti et al., 2022).

In Zarebski et al., 2022, we described an efficient and accurate method to approximate the likelihood of a point process of viral genomes and a time series of case counts, which we call Timtam. While Timtam resolved a long-standing challenge for the field, (i.e. how to reconcile genetic and classical epidemiological data in a computationally feasible way), it had some substantial limitations: a.) while efficient, it is a complicated algorithm lacking a convenient implementation, limiting reuse and making it inaccessible to most potential users; and b.) it only estimated prevalence at the present, not prevalence through time. Here we show how to resolve these outstanding problems. We resolve the first by implementing the approach in a BEAST2 package (Bouckaert et al., 2019) called Timtam, which is available on CBAN and can be installed via BEAUti. We resolve the second limitation with an extension to the algorithm, which enables conditioning on historical prevalence so it can be treated as a model parameter and estimated.

We carried out a simulation study to demonstrate that the methodology leads to well-calibrated estimates, i.e. that approximately 95% of the 95% HPD intervals (i.e. the credible intervals) contain the true parameter value from the simulation. We further demonstrate the “real-world” use of this package with two empirical case studies. In the first, we repeat an analysis by Andréoletti et al., 2022 of SARS-CoV-2 data from an outbreak on the Diamond Princess cruise ship. We infer a greater prevalence of infection than Andréoletti et al., 2022. We attribute the difference in prevalence to a limitation of the implementation in Andréoletti et al., 2022, which Timtam overcomes. In the second, we reanalyse data from the 2010 outbreak of poliomyelitis in Tajikistan from Yakovenko et al., 2014 and the Centers for Disease Control and Prevention (CDC), 2010 and compare the results to a similar analysis from Li et al., 2017.

## Methods

Figure 1A provides an example *transmission* tree, a complete description of who-infected-whom during an epidemic, along with the timing of these events, and the surveillance of this process. Sequenced cases appear as filled circles in the figure, unobserved cases end the lineage without a circle (at the time the case is unable to cause any onward transmissions, i.e. becomes uninfectious), and unfilled circles indicate scheduled observation of cases without sequencing, which occurred at the three times indicated with dashed lines. Figure 1B depicts the corresponding *reconstructed* tree and the time series of case counts. The reconstructed tree is the subtree of the transmission tree that results from pruning any leaves not corresponding to a sequenced sample. Unlike the transmission tree, the reconstructed tree is not oriented, consequently it does not specify which of the two descendant lineages of an internal node corresponds to the infector. The leaves of the transmission tree corresponding to unsequenced samples form a separate, but not independent, time series of confirmed cases. Figure 1C depicts the corresponding plots of the prevalence of infection through time (grey line) and the *lineages through time* (LTT) plot for the reconstructed tree (dashed line). The *lineages through time* (LTT) plot describes the number of lineages in the reconstructed tree as a function of time. Typically, the value of the LTT plot will be less than the prevalence of infection. In Figure 1C, a single estimate of the prevalence appears near the present as a star.

**Figure 1:**
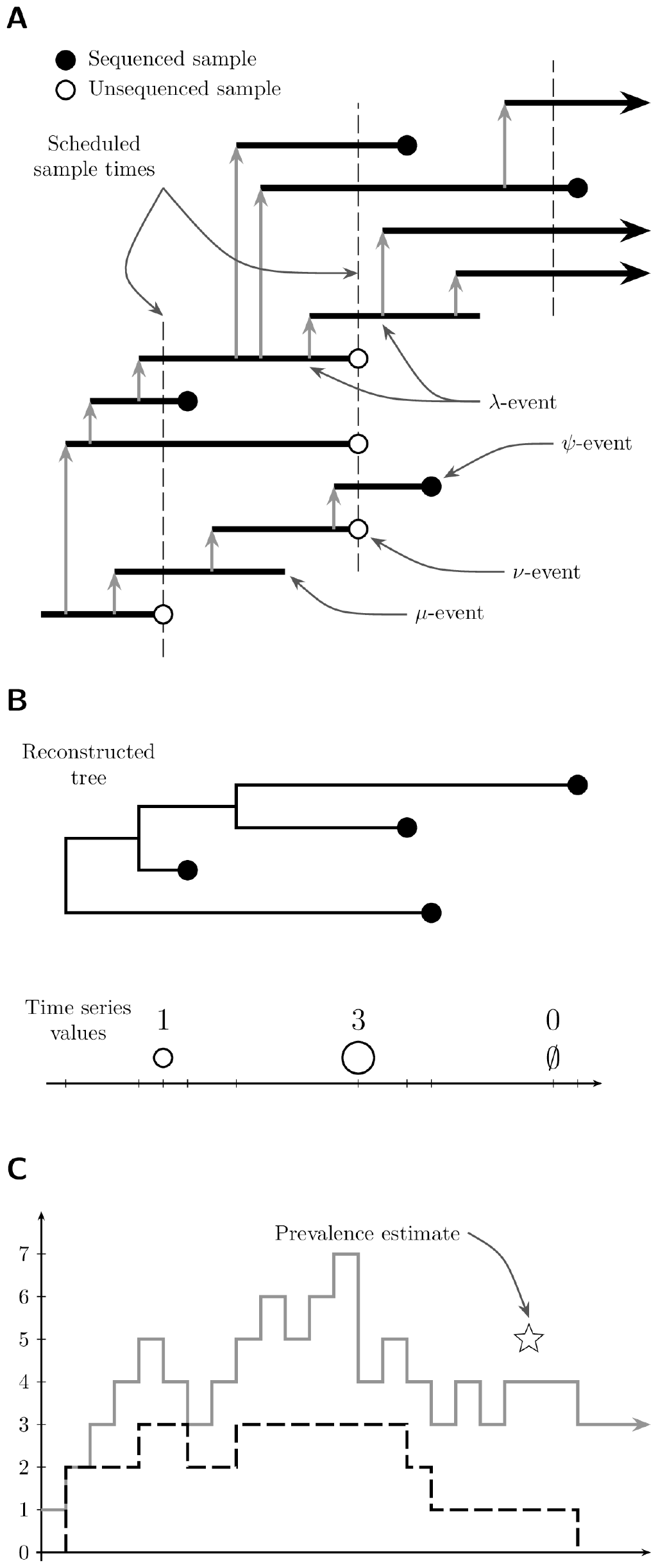
The transmission process is viewed as a sequence of events with the observations processed (in order) to approximate their joint likelihood. Panel **A** demonstrates a *transmission* tree with intervals of time an individual was infected indicated by horizontal lines and the vertical grey arrows indicating transmission. Three scheduled unsequenced samples are taken at the times indicated by the vertical dashed lines. Panel **B** shows the corresponding *reconstructed* tree and time series of confirmed cases in each of the scheduled unsequenced samples. In the third sample no cases were observed. Panel **C** shows the prevalence of infection (grey line) and the LTT (black dashed) along with a single (hypothetical) estimate of the prevalence (the star).

We refer to the lineages in the transmission tree that are not in the reconstructed tree as the *hidden* lineages because they are not visible in the raw data. At any given time the sum of the number of hidden lineages and the LTT of the reconstructed tree is equal to the prevalence of infection. We denote by *k*_*t*_ the value of the LTT at time *t* and by *H*_*t*_ the number of hidden lineages.

In the terminology of Zarebski et al., 2022, the reconstructed tree consists of sequenced unscheduled data, and the time series of cases represents unsequenced scheduled data. We may consider arbitrary combinations of (un)sequenced and (un)scheduled data, but here we focus on data sets that consist of sequenced unscheduled data and unsequenced scheduled data (i.e. time-stamped sequences and a time series of cases), since this aligns closest to typical epidemiological data sets.

In an epidemiological setting, we are often interested in the prevalence of infection and *ℛ*_e_(*t*), because these quantities are of critical importance when assessing the threat posed by an outbreak of infectious disease. Bayesian phylodynamic methods provide a coherent solution with clear quantification of uncertainty. Unfortunately, this usually requires us to evaluate the joint posterior distribution of the model parameters and the reconstructed tree (up to an unknown normalisation constant if we are using MCMC to generate posterior samples), conditioning on time-stamped viral genomes and a time series of confirmed cases. To do this, we need to evaluate the log-likelihood function in a computationally efficient way. Simulation based methods exist, but tend to be far more computationally expensive, which can reduce the utility of the resulting estimates if they are time-sensitive.

The data for our model consists of 𝒟_MSA_ and 𝒟_cases_, where 𝒟_MSA_ is the multiple sequence alignment (MSA) containing the time-stamped pathogen genomic data, and 𝒟_cases_ is the observation of confirmed cases without associated pathogen genomes.

The parameters of this process partition into four groups:

- ℋ, the number of hidden lineages at specified points in time (which we use to estimate the prevalence of infection);
- *𝒯*, the time-calibrated reconstructed tree describing the ancestral relationships between the sequences in 𝒟_MSA_;
- *θ*_evo_ the parameters of the evolutionary model, describing how genome sequences change over time (e.g. the molecular clock rate and nucleotide substitution model relative rate parameters);
- and *θ*_epi_, the parameters of the epidemiological model, describing how the outbreak/epidemic grows or declines over time and how we observe it.

Using the terminology of birth-death processes, *θ*_epi_ contains the birth rate *λ* and the death rate *μ* along with the sequenced sampling rate *ψ*, the unsequenced sampling rate *ω* (a.k.a. the occurrence rate), probability of observation in a scheduled sequenced sample *ρ* and the probability of observation in a scheduled unsequenced sample *ν*. Examples of these events are shown in Figure 1A. Throughout this manuscript, we treat these parameters as piecewise constant functions with known change times.

We can express the posterior distribution, *f* (ℋ, 𝒯, *θ*_epi_, *θ*_evo_ | 𝒟_MSA_, 𝒟_cases_), in terms of simpler components with the factorisation in Equation (1) below. The likelihood of the sequence data given the reconstructed tree and genomic parameters *f* (𝒟_MSA_ | 𝒯, *θ*_evo_), which appears in Equation (1), is sometimes called the *phylogenetic likelihood*. This function is well-known and can be efficiently calculated with Felsenstein’s pruning algorithm (Felsenstein, 1981). The likelihood of the time series of cases, reconstructed tree and prevalence, given the epidemiological parameters *f* (𝒟_cases_, 𝒯, ℋ | *θ*_epi_), often called the *tree prior*, is more accurately called the *phylodynamic likelihood*. Here we make the standard simplifying assumption that there is no dependence between the tree structure and the sequence evolutionary process. Consequently the phylogenetic likelihood is independent of 𝒟_cases_, ℋ and *θ*_epi_. We also assume *θ*_epi_ and *θ*_evo_ have independent priors.

We summarise the LTT and time series of unsequenced observations as a sequence of labelled events, each with an associated time in order to evaluate the phylodynamic likelihood *f* (𝒟_cases_, 𝒯, ℋ | *θ*_epi_). There are four types of events we consider:

1. new infections (births), corresponding to the internal nodes of the reconstructed tree;
2. unscheduled sequenced samples, corresponding to the leaves of the reconstructed tree;
3. scheduled unsequenced samples, corresponding to the elements of the time series of cases;
4. and pseudo-observations of the number of hidden lineages at prespecified times.

These events are not the same as the data: event types 1 and 4 are parameters. Event types 3 and 4 have a value associated with them: for a scheduled unsequenced datum this is the value of the time series, and for the pseudo-observations this is the number of hidden lineages. Note also that only a subset of all infection events in the epidemic will be captured by the reconstructed tree (see Figure 1). We denote the event observed at time *t*_*j*_ by 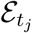, and the sequence of events that occur up until time *t* (inclusive) by ε _*≥t*_. We use *K*_*j*_ to indicate the value of the LTT of 𝒯 at time *t*_*j*_. In the following we order events using a backward time formulation, with 0 being the present (the time of the most recent sequenced sample in our data) and events further in the past having a larger time: *t*_0_ *> t*_1_ *> · · · > t*_*N*_. A consequence of specifying time in this way is that events occurring after the time of the last sequenced sample have negative times.

Expressions of the form 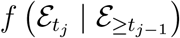 are simpler, so we will consider the following factorisation:

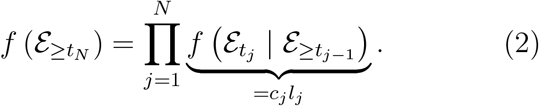

The factors in the product in Equation (2) are further divided into two parts: *c*_*j*_, which is the likelihood of the interval of time between *t*_*j−*1_ and *t*_*j*_ during which we observed nothing, and *l*_*j*_ the likelihood of the event that we observed at time *t*_*j*_. We start by considering *c*_*j*_. For *t*_*j−*1_ *> t > t*_*j*_ let 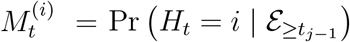, for *i ≥* 0 i.e. the joint distribution of the number of hidden lineages and not having observed any events since *t*_*j−*1_. Evaluated at *t*_*j*_ this becomes 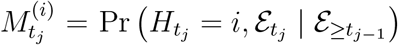i.e. the joint distribution of the number of hidden lineages and the event observed at time *t*_*j*_.

Consider the time during which we know there are no observed events, i.e. over an infinitesimal time step, *δt*, inside the interval *t*_*j−*1_ *> t > t*_*j*_. In this case, 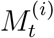 satisfies the following equation (up to leading order):

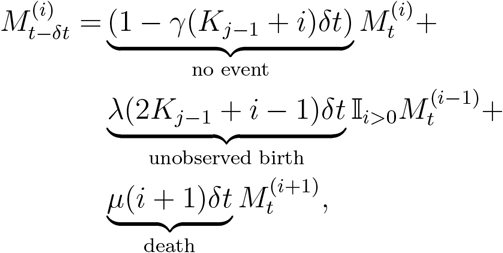

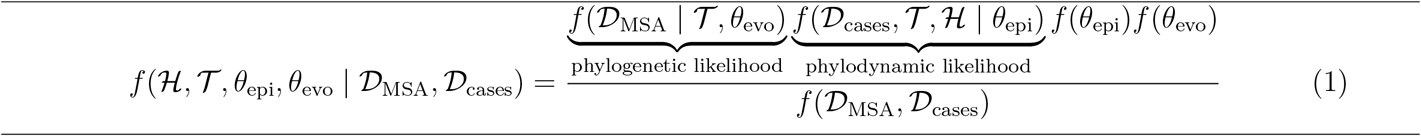

where 𝕀_*x*_ is the indicator random variable for event *x*. The factor of 2 in the term corresponding to unobserved births appears because the birth event creates two lineages (moving forward) and there are two ways to select one of them to be a hidden lineage and the other to continue the reconstructed tree.

Re-arranging the terms of these equations and taking the limit as the time step vanishes, we retrieve the master equations for this distribution, i.e. the system of differential equations that describe how it changes across the interval *t*_*j−*1_ *> t > t*_*j*_:

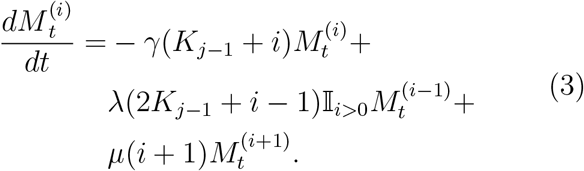

Let *M*_*t*_(*z*) be the generating function for this system of differential equations: 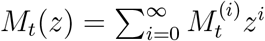.We can write the system in Equation (3) as the following PDE which has the generating function *M*_*t*_(*z*) as its solution.

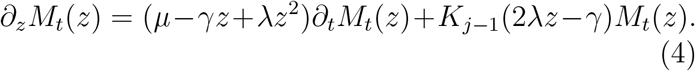

Manceau et al., 2020 solved Equation (4) in terms of results from Stadler, 2010. This partial differential equation allows us to update *M*_*t*_(*z*) across the intervals without observed events: (*t*_*j−*1_, *t*_*j*_).

Given the generating function across each interval we can evaluate the *c*_*j*_ in Equation (2), since 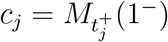. Note that we have to take the limit as time decreases to *t*_*j*_ because we are working with backwards time and there is a discontinuity.

The form of 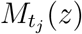 depends on both the limit 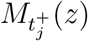 and the event observed at *t*_*j*_. To simplify the description below, let 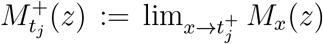 which is the limiting value of the generating function before the observation. How we transform the generating function depends on the type of 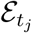. The expressions for *l*_*j*_ and 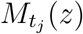 are as follows:

- for *λ* events *l*_*j*_ := *λ* and 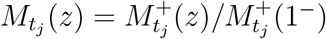;
- for *ψ* events *l*_*j*_ := *ψ* and 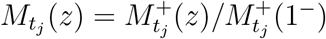;
- for *ω* events 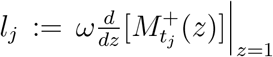 and 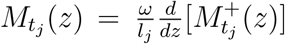;
- for *ρ* events when Δ*K*_*j*_ individuals were sampled and there are *K*_*j*_ lineages in the reconstructed tree just after the event, 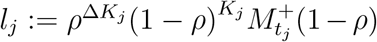 and 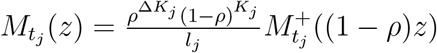;
- and for *ν* events when Δ*H*_*j*_ cases were observed, 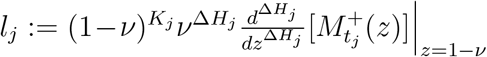 and 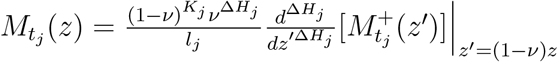

When co-estimating the prevalence, there is an additional event corresponding to a pseudo-observation of the number of hidden lineages. When we condition on 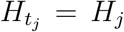, then 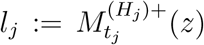 (which is the coefficient of 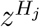 in 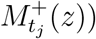 and 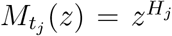, i.e. the generating function of the degenerate distribution corresponding to *H*_*j*_ hidden lineages. Note that while the ℋ_*j*_ are parameters of this model, we still include the corresponding *l*_*j*_ because we are also evaluating their prior distribution under the birth-death process. A novel contribution from this paper is the co-estimation of prevalence through time: the prevalence is treated as proper random variable under the posterior distribution.

### Timtam

We can compute the *c*_*j*_ analytically, however, we lack a closed form for the *l*_*j*_. In Zarebski et al., 2022, we describe the time series integration method through approximation of moments (Timtam). Timtam matches moments to approximate the relevant distribution with a negative binomial distribution. The generating function of the negative binomial distribution allows us to efficiently and accurately approximate the *l*_*j*_ and hence the likelihood of the model.

### The effective reproduction number

The reproduction number describes the average number of secondary infections: there are multiple ways to make this definition precise. We choose to define the effective reproduction number ℛ_e_(*t*) as the expected number of secondary infections generated by a newly infected individual from time *t* onward.

Without scheduled sampling, the value of ℛ_e_ in the birth-death model is *λ/*(*μ* + *ψ* + *ω*). Including scheduled sampling complicates matters because it combines continuous and discrete sampling. We derive a closed form expression for *ℛ*_e_ in the Supplementary Information. However the result is unwieldy so we will instead make use of a simple approximation.

### An approximation to ℛ_e_ for scheduled data

Consider the case of unscheduled sequenced and scheduled unsequenced samples at regular intervals of duration Δ_*t*_. From the perspective of an infectious individual, given they are removed during some scheduled sample, the number of intervals until this occurs, *W*, has a geometric distribution with probability *ν*. Given the scheduled samples occur at regular intervals of duration Δ_*t*_, the waiting time is approximately Δ_*t*_(*W* + 1*/*2) (where the half has come from a continuity correction). Provided Δ_*t*_ and *ν* are small, this distribution will be similar to an exponential distribution. The rate of an exponential distribution with the same mean is 2*ν/*(2Δ_*t*_ *− ν*Δ_*t*_).

This suggests the following approximation: we replace the scheduled unsequenced sampling (which complicate ℛ_e_ calculations) with unscheduled unsequenced sampling at rate 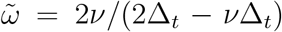(for which it is simple to calculate the ℛ_e_). I.e. we approximate the scheduled sampling with unscheduled sampling at a comparable rate, 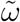, obtained by matching the geometric and exponential distributions as above. With unscheduled sampling at this rate, ℛ_e_ can be written as 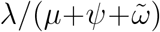. Figure 2 shows the effective reproduction number calculated using both the recursive method described in the Supplementary Information and the approximation that results from expressing the scheduled sampling as a rate 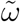. The values of ℛ_e_ are greater for longer intervals between scheduled samples, Δ_*t*_, because there is a longer duration during which the individual can infect others before being removed in a scheduled sample.

**Figure 2:**
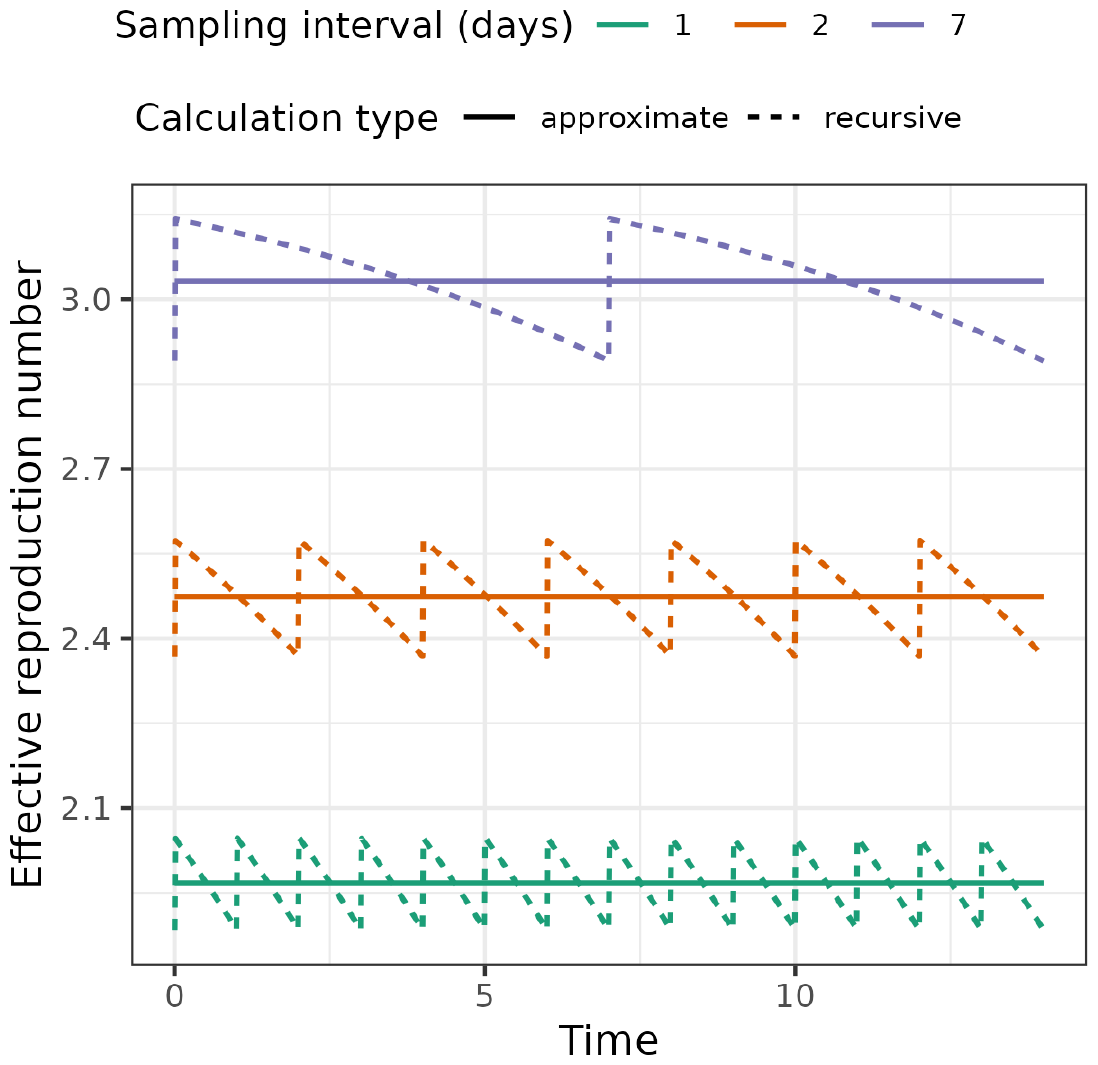
The approximation smooths out the saw-tooth value of the effective reproduction number in the case of scheduled samples. The parameters used for this figure are birth rate of 0.4, death rate of 0.1, sampling rate of 0.02 and a scheduled unsequenced sampling probability of 0.08 (at varying intervals). The solid lines indicate the values obtained with our approximation and the dashed lines indicate the true values accounting for scheduled sampling.

### Model parameterizations

There are multiple ways to parameterize this process. We refer to the parameterization in terms of the rates *λ, ψ, ω*, and probabilities *ρ*, and *ν* as the *canonical parameterization*. We derive the approximate likelihood in terms of these parameters. In an epidemiological context, with a time series of confirmed cases and point process sequence data, we prefer to parameterize the process in terms of the effective reproduction number, ℛ_e_, the net removal rate, 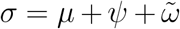 (where 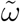 is as described above), and the observed proportion of infections captured by the time series, 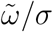, and the point process, *ψ/σ*, respectively. Given the focus on the use of time series data, we refer to this parameterization as the *time series parameterization*.

Note that when we use the time series parameterization in the SARS-CoV-2 and poliomyelitis analyses, we use the approximation of 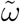 to simplify the model specification. This avoids the issue of having to adjust for future scheduled sampling in ℛ_e_(*t*).

### Sampled ancestors

A natural extension to this model is the inclusion of sampled ancestors (Gavryushkina et al., 2014), which Manceau et al., 2020 and Andréoletti et al., 2022 have already considered. Including sampled ancestors involves a probability *r* that an infected individual is removed upon (unscheduled) observation. Currently we assume all individuals are removed upon observation. We have not yet implemented this extension in Timtam, however we include the relevant expressions and the explanation of some additional details in the Supplementary Information, and leave the implementation as an exercise for the future.

## Results

### Calibration study

To assess the calibration of Timtam and the validity of our approximation of ℛ_e_, we carried out a simulation study. We sampled 100 epidemics from a birth-death process using remaster (Vaughan, 2024). Each epidemic ran for 56 days with the birth rate decreasing on day 42, (i.e. boom-bust dynamics), and two types of surveillance: sequenced and unsequenced with fixed rates, (see Table 1.) We assume a known removal (death) rate. The prevalence of infection in each of the simulated epidemics is shown in Fig. S1. There is a substantial amount of variability in the prevalence across the simulations, but the boom-bust dynamics can be seen in the average of the simulation trajectories.

**Table 1:** The simulated data for the calibration study was sampled from a birth-death process with two types of sampling and a change in birth rate leading to “boom-bust” dynamics. A final sequence sample is collected at the end of the simulation to ensure a consistent duration across the 100 replicates. Each simulation was conditioned to have at least two sequenced samples and a positive final prevalence of infection.

| Event | Rate | Transition |
| --- | --- | --- |
| Infection | $\lambda(t)$ | $X \xrightarrow{\lambda} 2X$ |
| Removal | $\mu = 0.046$ | $X \xrightarrow{\mu} \emptyset$ |
| Sequence | $\psi = 0.008$ | $X \xrightarrow{\psi} \text{Sequence}$ |
| Occurrence | $\omega = 0.046$ | $X \xrightarrow{\omega} \text{Case}$ |

We assume a known death rate, $\mu = 0.046$ . The number of infectious individuals, $X$ , was simulated for 56 days (after starting with a single infection $X(0) = 1$ ). The birth rate is $\lambda(t) = 0.185$ for $t < 42$ (“boom”: $\mathcal{R}_e = 1.85$ ) and $\lambda(t) = 0.0925$ for $t \geq 42$ (“bust”: $\mathcal{R}_e = 0.925$ ).

From each simulation we constructed two data sets: one with unsequenced samples treated as a point process, and a second with these samples aggregated into a time series of daily case counts. The parameters used are similar to those used in Zarebski et al., 2022 with an extension for the change in birth rate; we based the parameter values on the early dynamics of SARS-CoV-2 in Australia. The code implementing this simulation and the subsequent inference is available at https://github.com/aezarebski/timtam-calibration-study.

We sampled the posterior distribution of the model parameters for each simulated data set and compared the resulting estimates to the true values from the simulations. Figure 3 shows the estimates of prevalence and reproduction numbers across the simulations, ordered by the final prevalence in the simulation in the case where the unsequenced data is modelled as a point process. Figure 4 shows the corresponding results when the unsequenced data are aggregated into a daily time series of counts.

**Figure 3:**
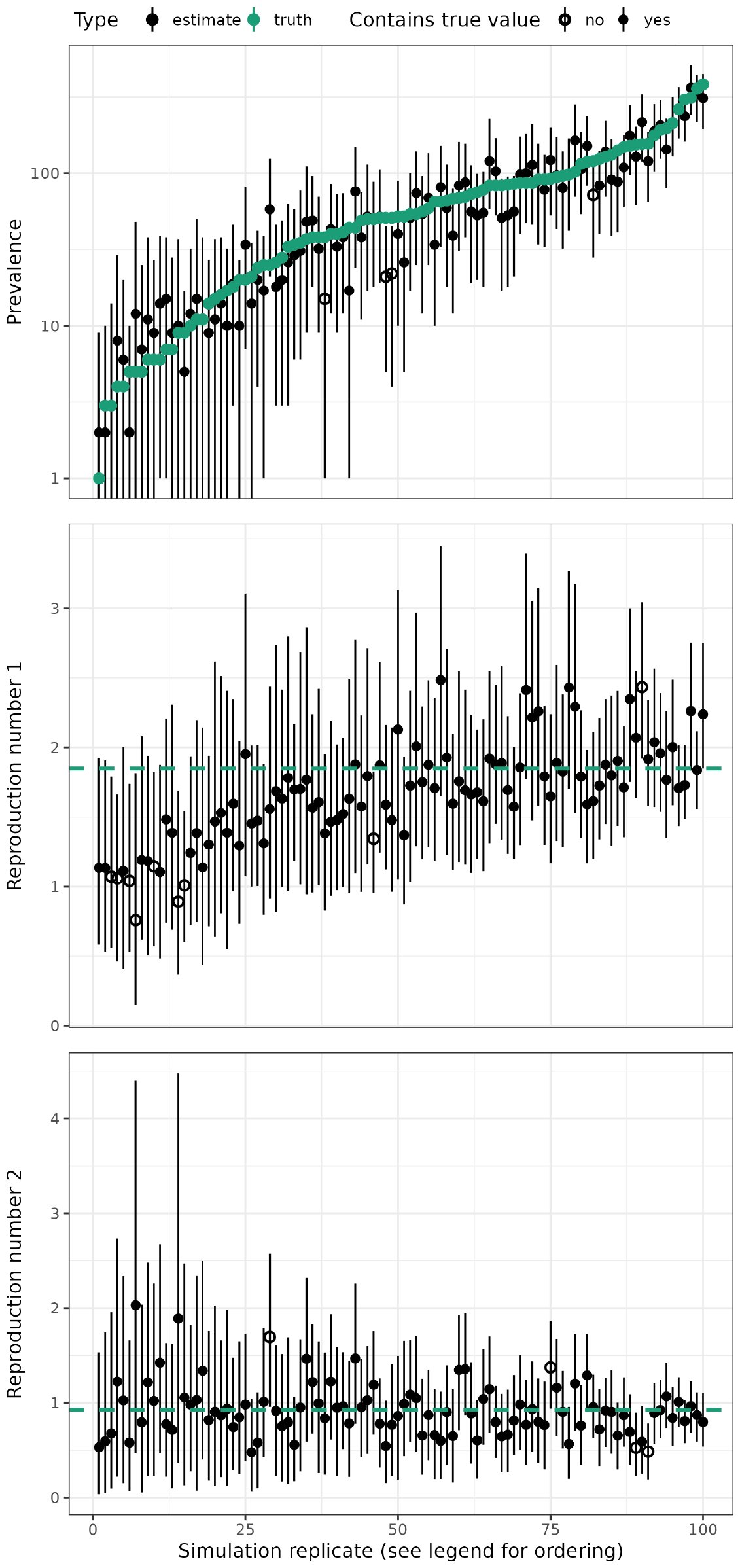
Parameter estimates converge to true values as the data set gets larger. The solid black lines display the HPD intervals, and points indicate the point estimates; the point is filled if the HPD interval contains the true value and empty if it does not. The green points and the green dashed lines indicate the true values of the final prevalence and the reproduction number in the boom and bust portions of the simulation. We ordered the replicates by the final prevalence in each simulation.

**Figure 4:**
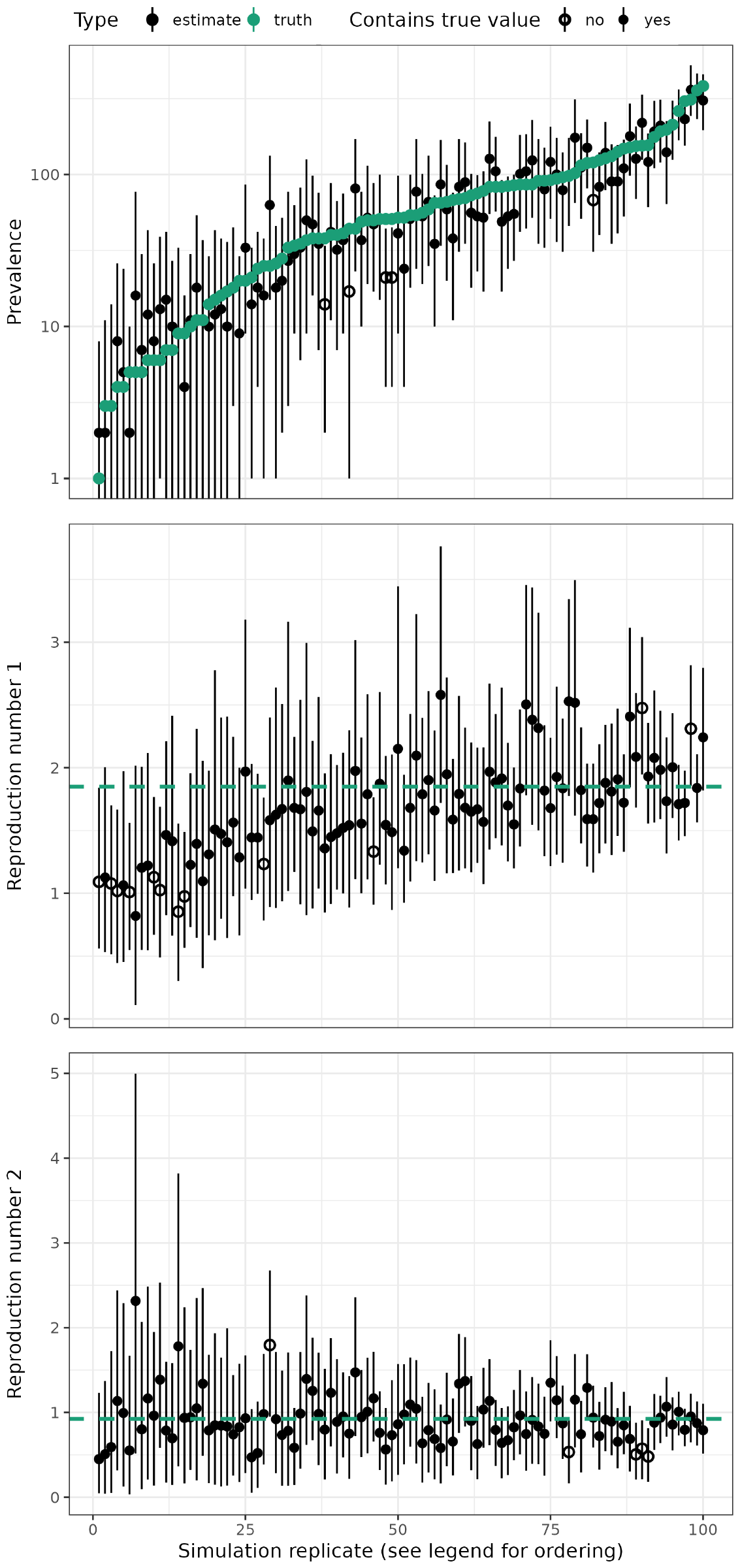
The estimated quantities and their true values from the simulation as shown in Figure 3 when the unsequenced observations were aggregated into a time series of daily case counts.

Comparing the simulations with a small final prevalence to those with a large final prevalence we see that for simulations with a larger prevalence the estimates of the reproduction number have less uncertainty and are less biased. We attribute this to the strong correlation between the final prevalence and the total number of data points (as shown in Fig. S2), as the reconstructed tree is likely to be larger for simulations with a larger final prevalence.

The credible interval is distinct from the confidence interval. (We have used the highest posterior density interval (HPD interval) as our credible interval, so have referred to them as “HPD intervals” rather than the less specific “credible interval”.) Due to the influence of the prior distribution, we do not necessarily expect 95% of the posterior distributions to contain the true parameters, however, we would like it to be close to this. We performed a hypothesis test of the null hypothesis that 95% of the HPD intervals contain the true parameter. Of course, the truth of this null depends upon the choice of prior distribution, nonetheless, we would like it to be difficult to falsify such a null hypothesis for plausible prior distributions.

In our hypothesis test, of the null hypothesis that the intervals are well-calibrated, we expect 91–99 of the HPD intervals to contain the true parameter value (out of the total 100 replicates). When interpreting the hypothesis test of whether the intervals are well calibrated at 95%, we need to bear in mind that the sampling of the limits of this interval are sparse (by construction) so one would ideally want a large effective sample size (ESS). Kruschke, 2014, p. 184 suggests an ESS *≥* 10000 is desirable for reliable 95% limits, while for each of our analyses the ESS was *≥* 200 for all variables.

Table 2 contains a summary of the rate parameter estimates from the first set of simulations (i.e. the ones with point process data) and Table 3 contains the corresponding summary for the second set of simulations (i.e. with unsequenced samples aggregated into a time series). For the estimates based on the point process data, the HPD intervals of both the reproduction number and the prevalence at the time of the last sequenced sample have a coverage that is consistent with the desired level (95%). This suggests the estimation method is well-calibrated. For the estimates based on the aggregated data, the coverage for 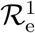 is lower than desired, however for both 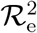 and *H* the coverage is suitable. This suggests that, despite the model misspecification due to aggregating the point process data, we are still able to generate good HPD intervals for the reproduction number and prevalence.

**Table 2:** Posterior parameter estimates and accuracy in the 100 simulations. There are boom-bust dynamics, for the first 42 days of the simulation the birth rate is *λ*_1_ after which it changes to *λ*_2_ for the subsequent 14 days. The death rate is assumed known.

| Par | True | Median | Error | Bias | Width | Coverage |
| --- | --- | --- | --- | --- | --- | --- |
| $\lambda_1$ | 0.185 | 0.186 | 0.116 | 0.004 | 0.523 | 94 |
| $\lambda_2$ | 0.092 | 0.095 | 0.337 | 0.032 | 1.375 | 94 |
| $\mu$ | 0.0460 | - | - | - | - | - |
| $\psi$ | 0.008 | 0.010 | 0.351 | 0.275 | 1.754 | 96 |
| $\omega$ | 0.046 | 0.052 | 0.248 | 0.140 | 1.163 | 98 |
| $\mathcal{R}_e^1$ | 1.850 | 1.689 | 0.180 | -0.087 | 0.664 | 91 |
| $\mathcal{R}_e^2$ | 0.925 | 0.897 | 0.291 | -0.030 | 1.132 | 96 |
| $H$ | - | - | 0.360 | -0.046 | - | 97 |

For each parameter (Par), the median over the 100 medians of the estimate, relative error, relative bias and the percentage of HPD intervals containing the true value is provided.

**Table 3:** Posterior parameter estimates and accuracy in the 100 simulations after we aggregated the unsequenced observations into daily counts and used the resulting time series as data.

| Par | True | Median | Error | Bias | Width | Coverage |
| --- | --- | --- | --- | --- | --- | --- |
| $\lambda_1$ | 0.185 | 0.186 | 0.121 | 0.003 | 0.535 | 91 |
| $\lambda_2$ | 0.092 | 0.094 | 0.337 | 0.018 | 1.377 | 95 |
| $\mu$ | 0.0460 | - | - | - | - | - |
| $\psi$ | 0.008 | 0.010 | 0.344 | 0.267 | 1.757 | 96 |
| $\tilde{\omega}$ | 0.046 | 0.053 | 0.265 | 0.143 | 1.185 | 98 |
| $\mathcal{R}_e^1$ | 1.850 | 1.670 | 0.191 | -0.097 | 0.655 | 88 |
| $\mathcal{R}_e^2$ | 0.925 | 0.873 | 0.287 | -0.057 | 1.141 | 95 |
| $H$ | - | - | 0.367 | -0.055 | - | 96 |

### SARS-CoV-2 on the Diamond Princess cruise ship

To demonstrate the utility of our new approach we replicated the analysis of a SARS-CoV-2 outbreak onboard the Diamond Princess cruise ship initially reported by Andréoletti et al., 2022. This outbreak is particularly well-suited to analysis because it occurred on an isolated cruise ship (with 3711 people onboard) in a carefully monitored population with detailed accounts of isolation and testing measures. The outbreak appears to have originated from a single introduction of the virus (Sekizuka et al., 2020). Figure 5 displays the cases and sequencing effort across the duration of the quarantine. We obtained a time series of daily confirmed cases from Dong et al., 2020 to use as 𝒟_cases_ and an alignment of 70 pathogen genomes (Sekizuka et al., 2020) was used as 𝒟_MSA_. The accession numbers for the sequences are available in the Supplementary Information.

**Figure 5:**
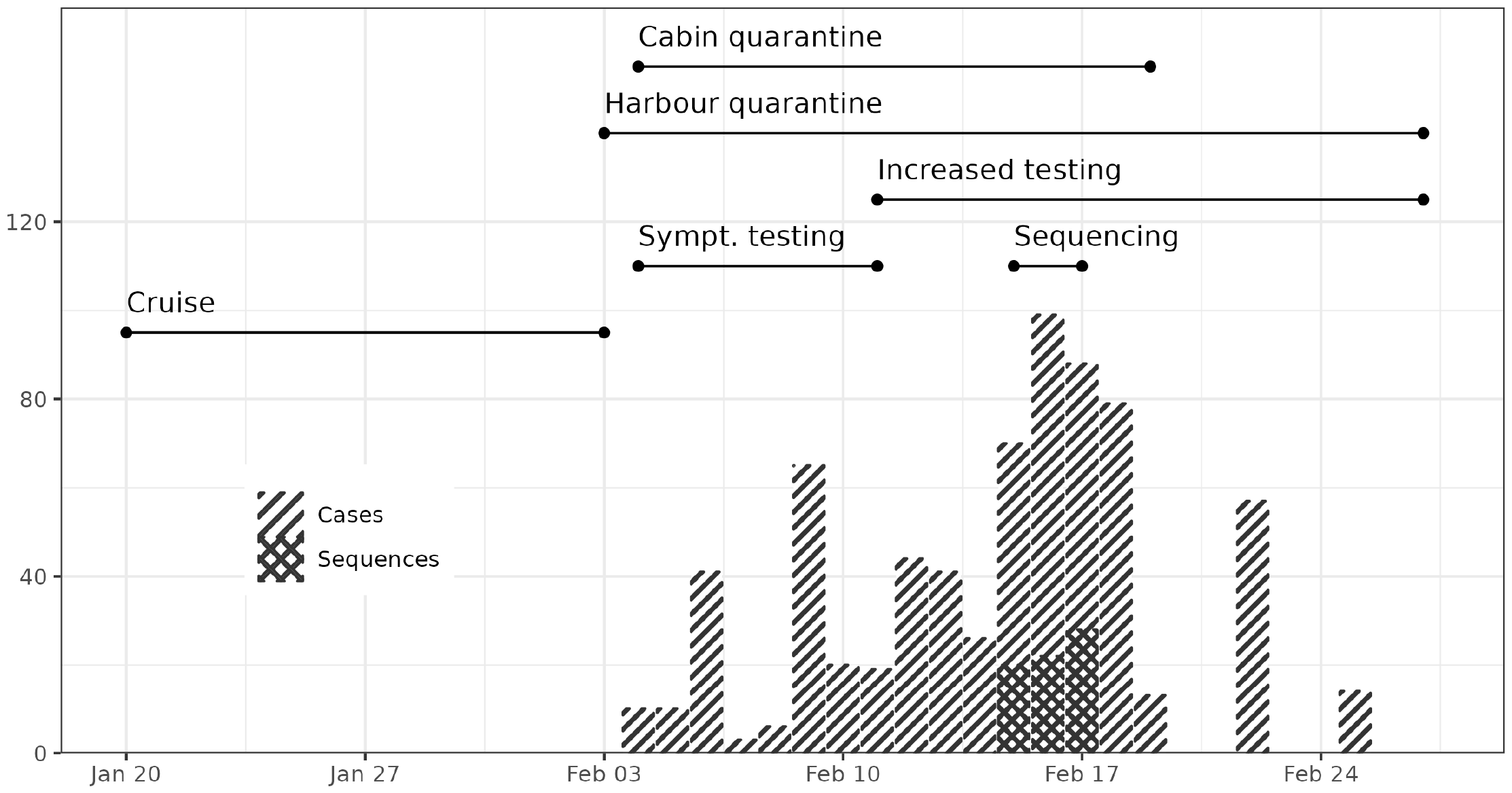
Sequences were collected across three days and testing varied throughout the quarantine period. The stacked bar chart shows the daily number of confirmed cases and sequenced samples. We indicate the timing of changes to surveillance and quarantine with lines at the top of the figure.

#### Model

We made minor adjustments to the model to better match standard epidemiological workflows for ℛ_e_ estimation, as described in the Supplementary Information. Importantly, we modelled daily case counts of confirmed cases as scheduled samples (i.e. a time series) instead of unscheduled samples (i.e. a point process of occurrences.) Additionally, we put an explicit prior on the reproduction number. Table S1 lists the prior distributions used in the model. The XML file specifying the analysis and post-processing are available from https://github.com/azwaans/timtam-diamond-princess.

#### Results

Figure 6 shows the estimates of the reproduction number through time along with the 95% HPD intervals. The estimates of the effective reproduction number are consistent with those from previous analyses of these data (Andréoletti et al., 2022, Figure 5). Our estimates differ from those of (Vaughan et al., 2024, Fig. S3). The discrepancy between the estimates from Vaughan et al., 2024 and ours may be due to the different data sets used: our analysis used both the time series of confirmed cases and pathogen genomes, while Vaughan et al., 2024’s estimates are based on genomic data alone.

**Figure 6:**
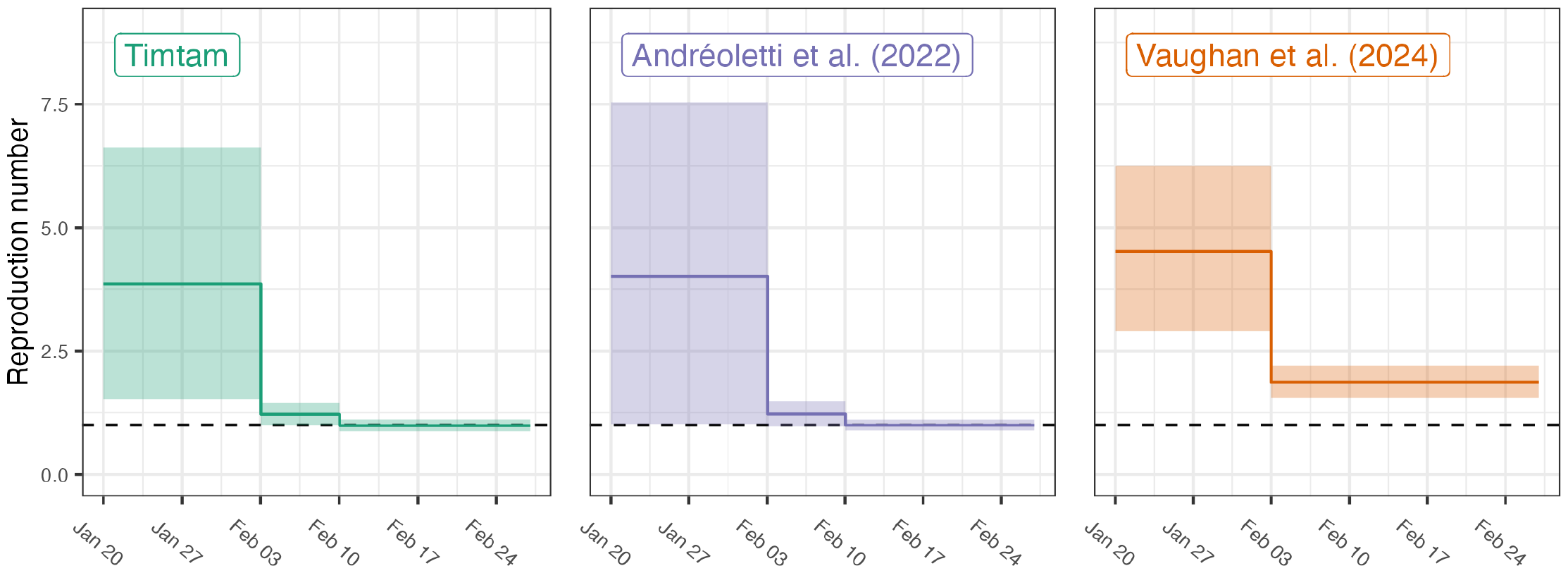
Estimates of the reproduction number and the 95% HPD intervals. In addition to our estimates (shown in green) estimates from Andréoletti et al., 2022 (purple) and Vaughan et al., 2024 (orange) are shown.

Figure 7 shows the estimates of the prevalence of infection and the 95% HPD intervals along with the corresponding values from Andréoletti et al., 2022. Our estimates suggest a larger prevalence of infection than the estimates from Andréoletti et al., 2022. Estimates from Vaughan et al., 2024 are not included as they estimated the cumulative number of infections, instead of the prevalence.

**Figure 7:**
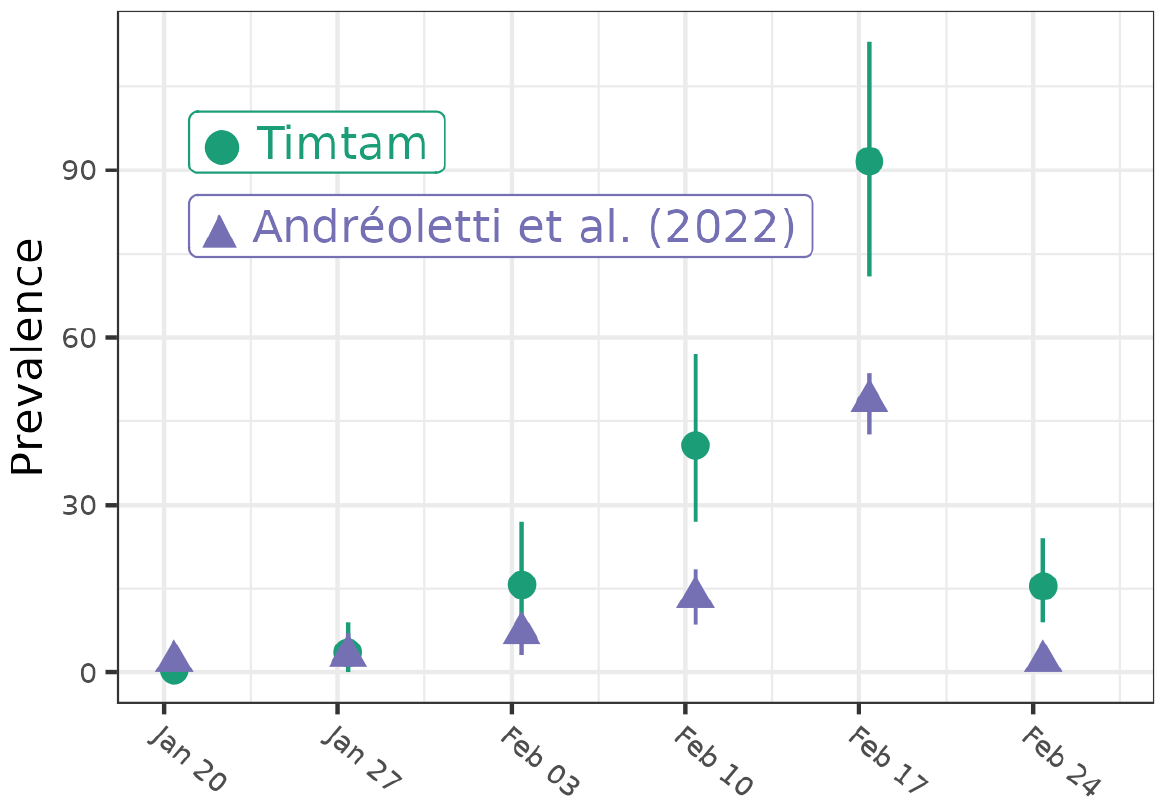
Estimates of the prevalence of infection and the 95% HPD intervals onboard the Diamond Princess. In addition to our estimates (green) estimates from Andréoletti et al., 2022 are shown (purple).

The MCMC chain to sample the posterior distribution (of both the tree and model parameters) ran in approximately an hour on a mid-range laptop. The effective sample size of each variable was *>* 300.

### Poliomyelitis in Tajikistan

Poliomyelitis, polio, is caused by infection with the poliovirus, an RNA virus spread through the fecal-oral route. While most poliovirus infections are asymptomatic, it has the potential to cause permanent paralysis. Polio has a long history but since the introduction of vaccines in the 1950s incidence has declined and there are sustained efforts towards eradication.

In 2010 there was an outbreak of wild poliovirus type 1 (WPV1) in Tajikistan. We re-analysed the genomic and time series data collected during the outbreak. These data had previously been jointly analysed by Li et al., 2017 using an age-structured model from Blake et al., 2014. A previous genomic analysis by Yakovenko et al., 2014 suggests the outbreak of WPV1 stemmed from a single importation in August–December 2009. However, substantial increases in the incidence of acute flaccid paralysis (AFP) did not occur until early 2010. A vaccination campaign was launched in May and the outbreak abated after that. Figure 8A shows a time series of cases and sequences generated; the timing of the rounds of vaccination is also shown.

**Figure 8:**
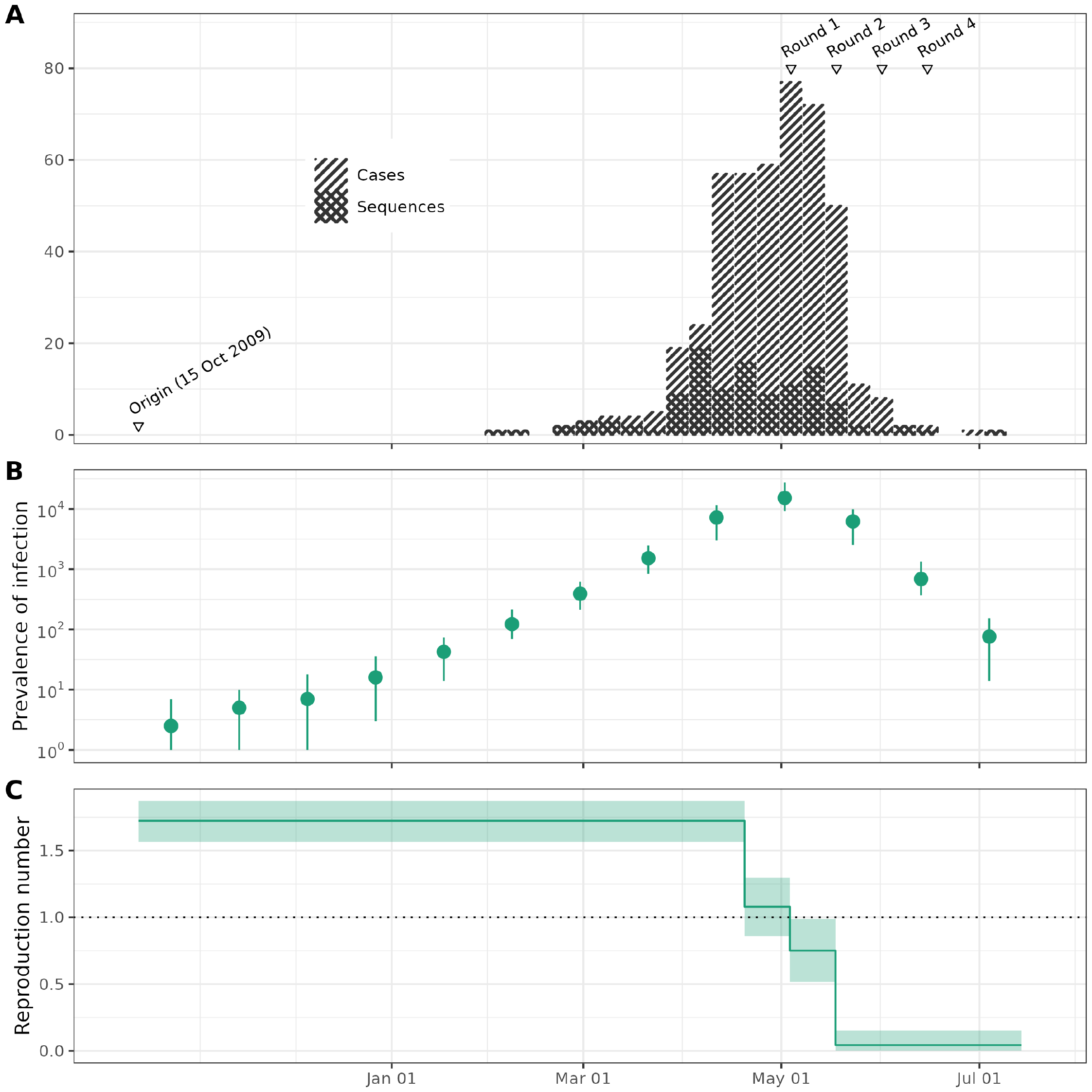
An outbreak of poliomyelitis in Tajikistan in 2010: **A**. Sequences were collected throughout the outbreak. The stacked bar chart shows the weekly number of confirmed cases and sequenced samples. We indicate the hypothesised origin time and the timing of vaccination rounds at the top of the figure. **B**. Estimates of the prevalence of infection (on a logarithmic scale) and the 95% HPD intervals at 30 day intervals across the outbreak. **C**. Estimates of the reproduction number and the 95% HPD intervals as constants before and after the start of vaccination.

#### Model

We modelled the transmission of poliovirus with a birth-death process with time varying effective reproduction number and surveillance rates to explain the effect of vaccination and heightened surveillance once the outbreak was recognised.

We extracted the weekly case counts of laboratory confirmed polio infections with paralysis as reported by the Centers for Disease Control and Prevention (CDC), 2010 (with WebPlotDigitiser (Rohatgi, 2021)) to use as 𝒟_cases_, and obtained an alignment of publicly available sequences from Li et al., 2017 (originally sequenced by Yakovenko et al., 2014) to use as 𝒟_MSA_. We subtracted the number of sequences from the time series to avoid duplication. As part of the sensitivity analysis we re-ran the analysis without this subtraction step and obtained similar estimates, (see Table S4.) Accession numbers for the sequences are available in the Supplementary Information.

Since case counts were only available at a weekly resolution, we distributed them uniformly across the days of the week and the sequenced samples uniformly within the date associated with them (when more than one genome was associated with the same date). I.e. cases were modelled as a daily time series of unsequenced samples and a point process of sequenced samples. Further details are available in the Supplementary Information and Table S2 lists the prior distributions used in the model. The XML files specifying the full analysis and post-processing are available from https://github.com/aezarebski/timtam-tajikistan.

#### Results

Figure 8B shows the estimates of the prevalence of infection and the 95% HPD intervals at 13 dates separated by 21 day intervals. Note that the estimates of the absolute prevalence extend before the first observed case. For example, we estimate that before February 2010 the prevalence was below 100. Even adjusting for a change in surveillance, there is little evidence of widespread transmission before February in the estimates of the prevalence of infection.

Figure 8C shows the estimates of the effective reproduction number through time along with the 95% HPD intervals. A full summary of parameter estimates can be found in Table S3 of the Supplementary Information. The estimates in Figure 8C suggest the effective reproduction number may have already started to decline before the beginning of the vaccination rounds, potentially due to public awareness. A comparison of these estimates with previous age-structured estimates is given in Figure S3.

The MCMC chain to sample the posterior distribution (of both the tree and model parameters) ran in approximately four days on a mid-range laptop and the effective sample size of each variable was *>* 200.

## Discussion

We implemented a model, which can also act as a phylodynamic tree prior to facilitate the co-estimation of the prevalence and the effective reproduction number. The resulting model can draw on both sampled pathogen sequence data and an epidemic time series of confirmed cases (i.e. observations of infection for which the pathogen genome was not sequenced). The algorithm used to compute the (approximate) log-likelihood is fast, requiring a number of steps linear in the number of sequences and length of the time series of unsequenced cases (Zarebski et al., 2022). The implementation is available as a BEAST2 package and tutorials on the usage of the package are bundled with the source code: https://github.com/aezarebski/timtam2.

We extended the method previously developed in Zarebski et al., 2022 to estimate historical prevalence, by explicitly modelling prevalence as a model parameter. This differs from several previous approaches, in which estimates of the prevalence come from intermediate steps in the likelihood calculation or from posthoc simulation. Treating the prevalence as a bona fide parameter also means we can incorporate additional data concerning prevalence into the analysis. For example, if survey data on infection in a random sample from the population was available for specific dates (e.g. from seroprevalence surveys) we could condition the model on this as additional data.

We performed a simulation study to demonstrate that the method is well-calibrated, i.e. that approximately 95% of the 95% HPD intervals do contain the true value. The simulation study also demonstrated that the performance of the method does not degrade substantially when we aggregated the occurrence data into a time series.

We used the validated method to replicate two analyses of limited single-source outbreaks. The first, carried out by Andréoletti et al., 2022, is of an outbreak of SARS-CoV-2 aboard the Diamond Princess cruise ship. The second, empirical analysis of the 2010 outbreak of poliomyelitis in Tajikistan, uses data from Yakovenko et al., 2014 and the Centers for Disease Control and Prevention (CDC), 2010.

The outbreak of SARS-CoV-2 aboard the Diamond Princess cruise ship was a relatively small, well-contained outbreak, for which where the majority of infections were ascertained. However, only a small number of sequenced samples exist, all dating from a period of only three days (Figure 5). Our estimates of the reproduction number (displayed in Figure 6) are consistent with the values from Andréoletti et al., 2022 and are broadly similar to those from Vaughan et al., 2024. However, Vaughan et al., 2024 only used genomic data, which may explain the difference in the *ℛ*_e_ estimates.

Our prevalence estimates are greater than those from Andréoletti et al., 2022 (Figure 7). We attribute this difference to their implementation having an upper limit of 40 on the number of hidden lineages, which was necessitated by the computational complexity of the numerical integration algorithm used to compute the likelihood. As such, their estimates should be interpreted as lower bounds on the prevalence and not absolute estimates. Timtam overcomes this limitation by a negative binomial approximation of the the number of hidden lineages, making it efficient at estimating large numbers of hidden lineages and applicable to real-world epidemic scenarios.

We modelled the sequenced SARS-CoV-2 infections as a point process, consistent with previous analyses of the data. Where multiple samples were available for a particular day, we uniformly spaced the sequenced samples across the day the samples were collected. A more nuanced analysis would have modelled these samples as scheduled sequenced samples, however this would make the resulting estimates harder to compare to previous results and complicate the interpretation.

The empirical analysis of the 2010 outbreak of poliomyelitis in Tajikistan uses data from Yakovenko et al., 2014 and the Centers for Disease Control and Prevention (CDC), 2010. This is a much larger outbreak over a period of months instead of weeks. Although a large proportion of the ascertained cases (all presenting with AFP) were sequenced across the entire reporting period of the outbreak (Figure 8A), we expect that most infections were not ascertained, since the majority of poliovirus infections are asymptomatic.

It is not possible to directly compare our estimates of the effective reproduction number of poliomyelitis in Tajikistan to previous work. Timtam does not support structured populations yet, so we were not able to obtain age-specific ℛ_e_ estimates such as those reported in Li et al., 2017. However, our estimates of the effective reproduction number during the central four weeks of the outbreak are similar to a demographically weighted average of the estimates from Li et al., 2017.

In addition, Timtam allows us to obtain estimates of the outbreak prevalence through time. Our estimates suggest more than a hundred asymptomatic infections for every AFP case, which is consistent with previous estimates (Nathanson et al., 2010). To the best of our knowledge, prevalence estimates for this outbreak have not been reported elsewhere.

Our implementation does not yet support the use of *sampled ancestors* (Gavryushkina et al., 2014). Extending the approximation to handle this case seems feasible, however there are substantial engineering challenges involved in implementing this in the BEAST2 platform. As the model and its implementation are useful without this extension we present it as is, and include the expressions required for including sampled ancestors in the Supplementary Information.

In summary, the Timtam package is an efficient implementation of our model within the BEAST2 framework, where it can be combined with a multitude of other model components. The model is suitable as a tree prior or demographic model for unstructured outbreaks and provides similar functionality to the model presented in Andréoletti et al., 2022, with the added advantages of being able to incorporate unsequenced cases (observations) as a time series, being able to condition on historical prevalence estimates and being efficient enough to handle large trees.

## Data Availability

All data produced are available online as described in the manuscript

https://github.com/aezarebski/timtam-tajikistan

https://github.com/azwaans/timtam-diamond-princess

## Acknowledgements

We thank Dr Timothy Vaughan for patiently answering many questions during the implementation of the Timtam package.

We thank Dr David Jorgensen for helpful comments on the analysis of the poliomyelitis data set.

AEZ, BG and OGP are supported by The Oxford Martin Programme on Pandemic Genomics. AZ is supported by the European Research Council (ERC) under the European Union’s Horizon 2020 research and innovation programme grant agreement no. 101001077.

