## Supplementary information for "Estimating epidemic dynamics with genomic and time series data"

#### Background

Stadler, 2010 derived the probability an individual infectious at time $t$ in the past gives rise to exactly zero or one observed event (e.g., they are either sequenced or appear as an occurrence). These probabilities are key to the calculations from Manceau et al., 2020, in which they solved a PDE for the probability generating function (PGF) of the distribution of hidden lineages.

The operations that need to be applied to the PGF to compute the likelihood are intractable, so in Zarebski et al., 2022 we constructed a negative binomial approximation to describe the number of descendent lineages of an infection. As shown by Kendall, 1948, each lineage has a number of descendent lineages which is over-dispersed for the Poisson distribution, so the sum of all of these is also over-dispersed. This means that we never run into problems with the mean being greater than the variance after conditioning on the population being a particular size at some point in time. Full details of the derivation of the negative binomial approximation are given in Zarebski et al., 2022.

#### Effective reproduction number

In this section we derive a closed form expression for the effective reproduction number, $\mathcal{R}_\text{e}$, in the birth-death process with scheduled sampling events. Let $Y_t(x)$ be the number of infections caused by an individual during the interval $(t, t+x)$, given they were already infectious at time $t$[1]. The effective reproduction number can be written as

$$\mathcal{R}_\text{e}(t) = \lim_{x\to\infty} \mathbb{E}\left[Y_t(x)\right].$$

In the standard birth-death process, if births (new infections) occur at rate $\lambda$ and deaths (cessation of infectiousness) at rate $\mu$, then $\mathcal{R}_\text{e} = \lambda/\mu$. From the perspective of an infectious individual in this process it does not matter if they cease to be infectious because of a $\mu$ (removal without sampling), $\psi$ (sequenced sampling) or $\omega$ (unsequenced sampling) event. The infectious individual only experiences a net birth rate, $\lambda$, a net death rate, $\mu' := \mu + \psi + \omega$, and a sequence of times, $t_j$ for $j = 0, 1, \cdots$, when they may be sampled with probability $p_j$ (i.e. the scheduled sampling times). Note that these rates may at a sequence of times, $s_j$, when either the net birth or death rates of the process change.

Let $T_\text{sched} = \min\{t_j, s_{j'}\}$ (for the appropriate indices $j$ and $j'$) be the time of the next scheduled event and $T_{\mu'}$ be the time at which the individual would cease to be infectious in the absence of scheduled sampling. Since $T_{\mu'}$ has an exponential distribution with rate $\mu'$, we can observe that $\Pr\left(T_{\mu'} < T_\text{sched}\right) = 1 - e^{-\mu'(T_\text{sched}-t)}$.

To compute $\mathcal{R}_\text{e}(t)$, we write $\mathbb{E}\left[Y_t(x)\right]$ as the sum of two potential outcomes, whether the infectious individual is removed before or after $T_\text{sched}$:

$$\begin{aligned}\mathbb{E}\left[Y_t(x)\right] = \\ &\mathbb{E}\left[Y_t(x) \mid T_{\mu'} < T_\text{sched}\right]\Pr\left(T_{\mu'} < T_\text{sched}\right) + \\ &\mathbb{E}\left[Y_t(x) \mid T_{\mu'} > T_\text{sched}\right]\Pr\left(T_{\mu'} > T_\text{sched}\right).\end{aligned}$$

To compute the first term in the equation, note that for $x \geq T_\text{sched} - t$, we have $\mathbb{E}\left[Y_t(x) \mid T_{\mu'} < T_\text{sched}\right] = \mathbb{E}\left[Y_t(T_\text{sched} - t) \mid T_{\mu'} < T_\text{sched}\right]$, because there can be no further infections after the individual has ceased to be infectious. Given $T_{\mu'}$ and $T_{\mu'} < T_\text{sched}$, the number of infections due to the individual in this period follows a Poisson process. Therefore,

[1] Some authors will require that the individual was newly infected at time $t$, but since the process is a birth-death process and has no memory we do not need to consider this.

$$\mathbb{E}\left[Y_t(x) \mid T_{\mu'} < T_{\text{sched}}\right] \Pr\left(T_{\mu'} < T_{\text{sched}}\right) = \left(1 - e^{-\mu'(T_{\text{sched}}-t)}\right) \int_0^x \lambda\tau \underbrace{\frac{\mu' e^{-\mu'\tau}}{1 - e^{-\mu'(T_{\text{sched}}-t)}}}_{*} d\tau$$

where $*$ is the conditional density of the time at which the individual is removed, $T_{\mu'}$. In the case $x = T_{\text{sched}} - t$ this is

$$\frac{\lambda}{\mu'}\left(1 - e^{-\mu'(T_{\text{sched}}-t)}\right) - \lambda(T_{\text{sched}} - t)e^{-\mu'(T_{\text{sched}}-t)}$$

which has a nice interpretation: the first term is the proportion of the total number of infections independent of the scheduled sampling and the second term corrects for the over-counting by removing the expected number if the infection persisted past the scheduled sample.

There are two expressions for $\mathbb{E}\left[Y_t(x) \mid \; T_{\mu'} > T_{\text{sched}}\right]$ depending on if $T_{\text{sched}} = t_j$ or $T_{\text{sched}} = s_{j'}$. For $T_{\text{sched}} = t_j$, assume that the probability of being observed (and hence being removed from the infectious class) as part of the scheduled sample is $p$. In this case

$$\begin{aligned} &\mathbb{E}\left[Y_t(x) \mid T_{\mu'} > T_{\text{sched}}\right] \\ =&\mathbb{E}\left[Y_t(T_{\text{sched}} - t) \mid T_{\mu'} > T_{\text{sched}}\right] + \\ &(1-p)\mathbb{E}\left[Y_{T_{\text{sched}}}(x - (T_{\text{sched}} - t))\right] \\ =&\lambda(T_{\text{sched}} - t) + (1-p)\mathbb{E}\left[Y_{T_{\text{sched}}}(x - (T_{\text{sched}} - t))\right] \end{aligned}$$

where the first term is the expected number of infections prior to the scheduled sample, and the second term is the expected number after it. In the related case where $T_{\text{sched}} = s_{j'}$, we have the same expression but with $p = 0$ and with an appropriate change in the parameters.

The expressions above give a recursive definition of the effective reproduction number. If we assume that there are only a finite number of scheduled samples and rate changes this recurrence will terminate. Even in the case of scheduled sampling continuing without end, we can use it to compute increasingly accurate values, and it is possible to use it to bound the true value with this approximation. As mentioned in the main text, we do not need to use this expression as there is a convenient and principled approximation in terms of $\tilde{\omega}$.

### Extension to sampled ancestors

#### Observation with possible removal

Consider the situation in which, upon observation, there is a probability $r$ that the observed individual is removed from the infectious population, otherwise they remain in the infectious population and can continue to infect other individuals. Since this situation can lead to individuals who are observed but have sequenced descendants, we refer to them as *sampled ancestors*.

Since the PDE for the generating function in the likelihood assumes there are no observations, this portion of the likelihood remains the same. The only expressions that change are those involved with adjusting the generating function for $\psi$, $\omega$, $\rho$ or $\nu$ type observations.

Note that the potential for non-removal complicates sequenced samples. If $r = 1$, then every sequenced sample corresponds to a leaf in the reconstructed tree (a degree-one node). If $r < 1$, then there are two possibilities. The first is a degree-two node, a so-called *sampled ancestor*, which occurs when a sequenced individual is either sequenced again at a later date or has a sequenced descendant. The second, is a leaf with a label to indicate that it was not removed upon sampling but possibly gave rise to subsequent infections or unsequenced observations.

For unscheduled data, the expressions are the same as those already given by Manceau et al., 2020, however we will repeat them here to make it clear how they fit into our methodology. The relevant expressions for $l_j$ and $M_{t_j}(z)$ for the unscheduled observations, accounting for the possibility of non-removal, are as follows:

- for $\psi$ events with removal $l_j := \psi r$ and $M_{t_j}(z) = M^+_{t_j}(z)/M^+_{t_j}(1^-)$,
- for $\psi$ events without removal (with sequenced descendents) $l_j := \psi(1 - r)$ and $M_{t_j}(z) = M^+_{t_j}(z)/M^+_{t_j}(1^-)$,
- for $\psi$ events without removal (without sequenced descendents) $l_j := \psi(1 - r)$ and $M_{t_j}(z) = zM^+_{t_j}(z)/M^+_{t_j}(1^-)$,
- for $\omega$ events with removal $l_j := \omega r \frac{d}{dz}[M^+_{t_j}(z)]\Big|_{z=1}$ and $M_{t_j}(z) = (\omega r/l_j)\frac{d}{dz}[M^+_{t_j}(z)]$,
- and for $\omega$ events without removal $l_j := \omega(1 - r)z\frac{d}{dz}[M^+_{t_j}(z)]\Big|_{z=1} + \omega(1-r)kM^+_{t_j}(1)$ and $M_{t_j}(z) = (\omega(1-r)/l_j)\left\{z\frac{d}{dz}[M^+_{t_j}(z)] + kM^+_{t_j}(z)\right\}$,

The case of scheduled observations is complicated by the fact that we need to specify how removal works when multiple individuals have been observed. This differs from previous work on the problem in which a single $\rho$-sample at the present was considered in which case no distinction was needed.

We will assume that for sequenced samples we know the removal status of each individual, and for unsequenced samples we know the total number that where removed during a scheduled sample. Under these assumptions we have been able to derive the relevant expressions for $\rho$-sampling, but not $\nu$-sampling (in which case we have only been able to derive the expressions for the special cases of $r = 0$ or 1).

- For a $\rho$ event at time $t_j$ in which there are $\Delta K_j^{1-r}$ leaves without removal observed, $\Delta K_j^r$ leaves with removal observed, $A_j$ sampled ancestors observed, and $K_j$ lineages in the reconstructed tree just after the event, then, $l_j = r^{\Delta K_j^r}(1-r)^{\Delta K_j^{1-r}+A_j}\rho^{\Delta K_j^r+\Delta K_j^{1-r}+A_j}(1-\rho)^{K_j-A_j}M_{t_j}^+(1-\rho)$ and $M_{t_j}(z) = M_{t_j}^+((1-\rho)z)/M_{t_j}^+(1-\rho)$,
- and for a $\nu$ event at time $t_j$ in which there are $\Delta H_j$ cases observed, if all cases are removed, $r = 1$, and we can re-use the expressions from the main text, and if no cases are removed, $r = 0$, and $l_j = \nu^{\Delta H_j}\frac{d^{\Delta H_j}}{dz^{\Delta H_j}}\left[z^{K_j}M_{t_j}^+(z)\right]\Big|_{z=1-\nu}$ and $M_{t_j}^+(z) = l_j^{-1}(z\nu)^{\Delta H_j}\frac{d^{\Delta H_j}}{dz'^{\Delta H_j}}\left[z'^{K_j}M_{t_j}^+(z')\right]\Big|_{z'=(1-\nu)z} z^{-K_j}$.

### Calibration study

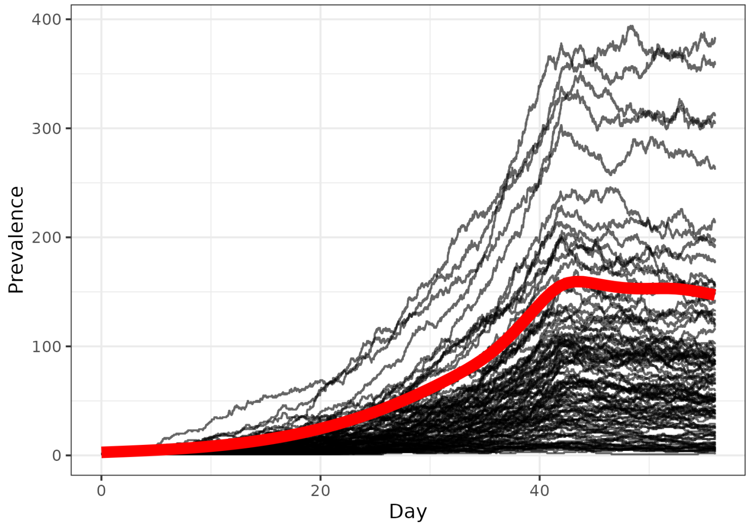


**Figure S1:** The number of infected individuals in each simulation. There is substantial variability between the prevalence of infection in the simulated epidemics (each displayed with a transparent black line), but the boom-bust dynamics are evident from the smoothed values (the solid red line).

Fig. S1 displays the trajectory of the prevalence in each of the simulated epidemics along with their average prevalence. In the main text we displayed the results of the calibration simulation study with the simulations ordered by their final prevalence. The posterior uncertainty in the parameter estimates appears to be smaller for simulations with a greater final prevalence. We explain this by noting that the final prevalence is strongly correlated with the total number of confirmed cases, and hence the amount of data available to inform the posterior distribution. Fig. S2 shows the correlation between the total number of confirmed cases and the final prevalence in each of the simulated epidemics used in the calibration study.

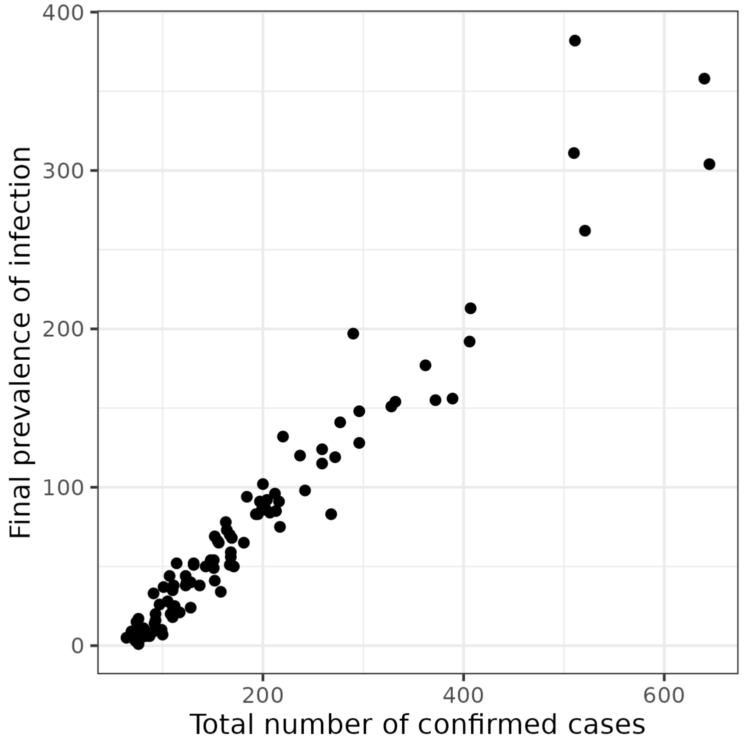


**Figure S2:** There is a strong correlation between the final prevalence of infection in the simulated epidemics and the total number of data points, (measured as total number of observed cases).

### SARS-CoV-2 on the Diamond Princess cruise

We based our analysis of the Diamond Princess cruise outbreak on the model from Andréoletti et al., 2022. Our analysis was implemented in BEAST2 (Bouckaert et al., 2019) using the Timtam package. The code for this analysis, the XML files and scripts for post-processing, are available at `https://github.com/azwaans/timtam-diamond-princess`. The MCMC chain ran in approximately an hour on a mid-range laptop and the effective sample size of each variable was $> 300$.

#### Data

We used an alignment of 70 SARS-CoV-2 sequences that were generated by Sekizuka et al., 2020. One sequence (of the 71) from the initial analysis by Andréoletti et al., 2022 was not publicly available and hence was included as an unsequenced case in the time series instead. The sequence accession numbers are given below:

EPI_ISL_416565,EPI_ISL_416566,EPI_ISL_416567,
EPI_ISL_416568,EPI_ISL_416569,EPI_ISL_416570,
EPI_ISL_416571,EPI_ISL_416572,EPI_ISL_416573,
EPI_ISL_416574,EPI_ISL_416575,EPI_ISL_416576,
EPI_ISL_416577,EPI_ISL_416578,EPI_ISL_416579,
EPI_ISL_416580,EPI_ISL_416581,EPI_ISL_416582,
EPI_ISL_416583,EPI_ISL_416584,EPI_ISL_416585,
EPI_ISL_416586,EPI_ISL_416587,EPI_ISL_416588,
EPI_ISL_416589,EPI_ISL_416590,EPI_ISL_416591,
EPI_ISL_416592,EPI_ISL_416593,EPI_ISL_416594,
EPI_ISL_416595,EPI_ISL_416596,EPI_ISL_416597,
EPI_ISL_416598,EPI_ISL_416599,EPI_ISL_416600,
EPI_ISL_416601,EPI_ISL_416602,EPI_ISL_416603,
EPI_ISL_416604,EPI_ISL_416605,EPI_ISL_416606,
EPI_ISL_416607,EPI_ISL_416608,EPI_ISL_416609,
EPI_ISL_416610,EPI_ISL_416611,EPI_ISL_416612,
EPI_ISL_416613,EPI_ISL_416614,EPI_ISL_416615,
EPI_ISL_416616,EPI_ISL_416617,EPI_ISL_416618,
EPI_ISL_416619,EPI_ISL_416620,EPI_ISL_416621,
EPI_ISL_416622,EPI_ISL_416623,EPI_ISL_416624,
EPI_ISL_416625,EPI_ISL_416626,EPI_ISL_416627,
EPI_ISL_416628,EPI_ISL_416629,EPI_ISL_416630,
EPI_ISL_416631,EPI_ISL_416632,EPI_ISL_416633,
EPI_ISL_416634

We gratefully acknowledge the following authors from the originating laboratories responsible for obtaining the specimens, as well as the submitting laboratories where the genome data were generated and shared via GISAID, on which this research is based. All submitters of data may be contacted directly via www.gisaid.org.

##### Originating Laboratory

Japanese Quarantine Stations

##### Submitting Laboratory

Pathogen Genomics Center, National Institute of Infectious Diseases

##### Authors

Tsuyoshi Sekizuka, Kentaro Itokawa, Rina Tanaka, Masanori Hashino, Tsutomu Kageyama, Shinji Saito, Ikuyo Takayama, Hideki Hasegawa, Takuri Takahashi, Hajime Kamiya, Takuya Yamagishi, Motoi Suzuki, Takaji Wakita, Makoto Kuroda

#### Genetic likelihood and associated priors

##### Site and clock model

We use an HKY substitution model with a strict clock and a fixed clock rate of $2.19178 \times 10^{-6}$ substitutions/site/day. This is equivalent to a rate of 0.0008 substitutions/site/year

##### Epidemiology and tree distribution

We initialised the analysis with a random starting tree and the tree is estimated as a model parameter in order to take phylogenetic uncertainty into account. Table S1 shows the prior distribution used for the parameters related to the tree likelihood.

The timeline of events and their epidemiological significance are outlined in Fig. 5 of the main text. The origin time of the outbreak is assumed to be 28.9 days prior to the present (the time of the last sequenced sample). In calendar time this is the 20th of January.

The birth rate $\lambda$ is assumed to change once on the 4th of February (14 days prior to the present), when the outbreak was discovered and interventions started. The net becoming uninfectious rate $\sigma$ is assumed to change on the 4th and the 11th of February, due to changes in surveillance. We assume a known natural becoming uninfectious rate (the "death rate" of the birth-death process) $\mu$ of 0.05 days$^{-1}$. This corresponds to an average infectious duration of 20 days (in the absence of any interventions).

We assumed a scheduled unsequenced sample was taken at midday every day from the 20th of January to the 27th of February, inclusive (note that four of these scheduled sampling events failed to collect any positive samples). The approximate proportion of removals appearing in the time series $p_{\tilde{\omega}} := \tilde{\omega}/\sigma$ is assumed to be zero prior to the 4th of February (start of symptomatic testing) and after that is assumed to change once on the 11th of February (increased testing).

The proportion of removals that are sequenced $p_\psi := \psi/\sigma$ is assumed to be non-zero only during the time period when sequenced samples were actually collected, i.e. from the start of the 15th to the end of the 17th of February. Sequenced samples are uniformly distributed *within* the day on which they were collected (assumed to be the date associated with the sequence on GISAID). To avoid conflicts due to multiple events occurring at the same time we (i) schedule all parameter change times at midnight, and (ii) if there are an odd number of sequenced samples on a day, the times are shifted slightly, so that none fall exactly on midday.

#### Prevalence estimation

We estimated the prevalence onboard each week, starting on the second week of the outbreak (as shown in Fig. 7 of the main text). To avoid any conflicts with parameter change times or sample times, these estimates refer to the prevalence at 9 am on each day of interest (i.e. date + 0.375).

| Parameter | Date | Prior |
|---|---|---|
| $H$ | Jan 20–Feb 24 (weekly) | by process |
| $\mathcal{R}_e$ | Jan 20–Feb 3 | lognormal(0.8, 0.5) |
| $\mathcal{R}_e$ | Feb 4–Feb 10 | lognormal(0.8, 0.5) |
| $\mathcal{R}_e$ | Feb 11–Feb 24 | lognormal(0.8, 0.5) |
| $\sigma$ | Jan 20–Feb 3 | 0.05 |
| $\sigma$ | Feb 4–Feb 10 | lognormal(-4.08, 1.0) |
| $\sigma$ | Feb 11–Feb 24 | lognormal(-2.73, 1.0) |
| $\psi/\sigma$ | Jan 20–Feb 14 | 0 (fixed) |
| $\psi/\sigma$ | Feb 15–Feb 17 | 0.214 (fixed) |
| $\psi/\sigma$ | Feb 18–Feb 24 | 0 (fixed) |
| $\tilde{\omega}/\sigma$ | Jan 20–Feb 3 | 0 (fixed) |
| $\tilde{\omega}/\sigma$ | Feb 4–Feb 10 | 0.786 (fixed) |
| $\tilde{\omega}/\sigma$ | Feb 11–Feb 24 | 1.0 (fixed) |

**Table S1:** Prior distributions for the model parameters used in the analysis of the Diamond Princess dataset with the time series parameterization.

### Poliomyelitis in Tajikistan

The analysis was implemented in BEAST2 (Bouckaert et al., 2019) using the Timtam package. The code for this analysis, the XML files and scripts for post-processing, are available at `https://github.com/aezarebski/timtam-tajikistan`. The MCMC chain ran in approximately four days on a mid-range laptop and the effective sample size of each variable was $> 200$.

#### Data

We used an alignment of 116 sequences (GenBank IDs: KC880365–KC880521), originally generated and submitted by Yakovenko et al., 2014, as aligned by Li et al., 2017. Each sequence is associated with a collection date.

We gratefully acknowledge the following authors from the originating laboratories responsible for obtaining the specimens, as well as the submitting laboratories where the genome data were generated: Yakovenko, M.L., Gmyl, A.P., Ivanova, O.E., Eremeeva, T.P., Ivanov, A.P., Prostova, M.A., Baykova, O.Y., Isaeva, O.V., Gavrilin, E.V., Lipskaya, G.Y., Shakaryan, A.K., Kew, O.M., Deshpande, J.M. and Agol, V.I.

#### Genetic likelihood and priors

##### Site and clock model

Following Li et al., 2017, we use a K80+$\Gamma$ substitution model (implemented by the SSM package (Bouckaert et al., 2017) with two rate categories) with a strict clock and a fixed clock rate of 0.00002740 substitutions/site/day. This is equivalent to 0.01 substitutions/site/year.

##### Epidemiology and tree distribution

As with the Diamond Princess dataset we again initialised the analysis with a random starting tree and the tree is estimated as a model parameter. Table S2 shows the prior distributions used for the parameters related to the tree likelihood.

Figure 8A shows the timing of the events and their epidemiological significance: the origin time of the outbreak is assumed to be 15 October 2009 (262 days before the last sequence was collected on 4 July 2010); we assume that prior to the first sequence on 1 February 2010 (153 days before the last sequence) the probability of sequencing and observation was zero; we assume the effective reproduction number changed three times, at 61 days prior to the collection date of the last sequence (when the first round of vaccinations began on 4 May) as well as two weeks before and after that date (to allow for changes in behaviour due to awareness of the outbreak and depletion of susceptibles due to continuing vaccination).

We chose the prior distribution on $\mathcal{R}_e$ and the rate of becoming uninfectious to reflect a broad range of plausible values (Blake et al., 2014). Since there is approximately 1 case of poliomyelitis in $\approx 200$ primary infections (Blake et al., 2014)[2], and the ratio of sequences to time series cases is approximately 1-to-3 we use the two beta priors in the Table S2 which have expected values that sum to 1/200 and have a ratio of 1-to-3.

| Parameter | Date | Prior |
|---|---|---|
| $H$ | Dec 6–Jul 4 (30 day intervals) | by process |
| $\mathcal{R}_e$ | All intervals | Normal$(2.0, 2.0^2)$ |
| $\sigma$ | | Uniform$(0.1, 1.0)$ |
| $\psi/\sigma$ | before Feb 1 | 0 (fixed) |
| $\psi/\sigma$ | from Feb 1 | Beta$(2, 1598)$ |
| $\tilde{\omega}/\sigma$ | before Feb 1 | 0 (fixed) |
| $\tilde{\omega}/\sigma$ | from Feb 1 | Beta$(3, 797)$ |

**Table S2:** Prior distributions for the model used in the analysis of the polio dataset from the 2010 outbreak in Tajikistan with the time series parameterization.

#### Results

Table S3 contains the point estimates and HPD intervals for the epidemiological parameters: $\mathcal{R}_e$, $\sigma$, $\psi$ and $\omega$. Figure S3 shows a comparison of the estimated effective reproduction through time against the estimates from Li et al., 2017 of the basic reproduction number in people aged $\leq 5$ and $> 5$ years of age along with a population weighted average of these values computed from the number of people in Tajikistan in each of these age

[2] Centers for Disease Control and Prevention (CDC), 2021 suggests that less than 1% of polio infections result in paralysis.

ranges in 2013 (1,287,331 and 6,786,935 respectively,) as reported by the UN (United Nations Statistics Division, 2013).

| Parameter | Estimate (95% HPD interval) |
|---|---|
| $\mathcal{R}_e$ (before 20 April) | $1.72\ (1.57, 1.87)$ |
| $\mathcal{R}_e$ (20 April – 4 May) | $1.08\ (0.86, 1.30)$ |
| $\mathcal{R}_e$ (4 May – 18 May) | $0.75\ (0.52, 0.99)$ |
| $\mathcal{R}_e$ (after 18 May) | $0.04\ (4.08 \times 10^{-7}, 0.15)$ |
| $\sigma$ | $0.11\ (0.10, 0.13)$ |
| $\psi/\sigma$ | $1.58 \times 10^{-3}(7.03 \times 10^{-4}, 2.63 \times 10^{-3})$ |
| $\tilde{\omega}/\sigma$ | $4.71 \times 10^{-3}(2.20 \times 10^{-3}, 7.82 \times 10^{-3})$ |

**Table S3:** Point estimates of epidemiological parameters (median value) and 95% HPD intervals.

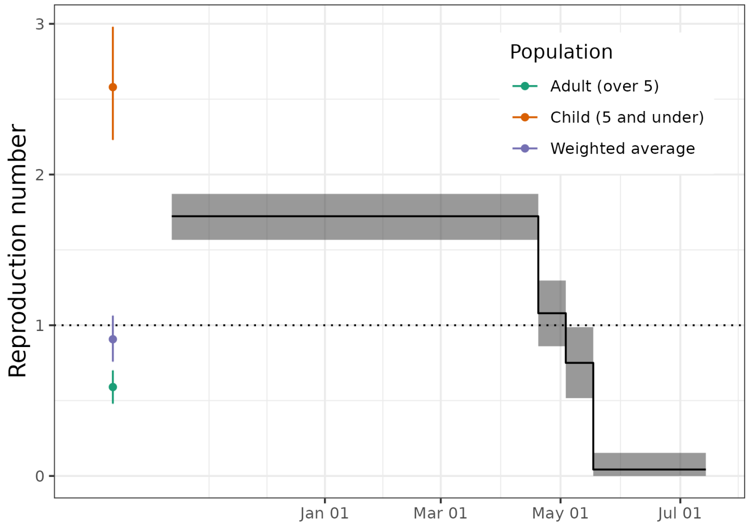


**Figure S3:** The posterior distribution of the effective reproduction number, $\mathcal{R}_e$, before and after May 4 along with estimates from Li et al., 2017 of the basic reproduction number, $\mathcal{R}_0$, in people aged $\leq 5$ and $> 5$ years of age along with a population weighted average of these values.

#### Marginal posterior/prior distributions

Figures S4, S5, S6, and S7 show the prior and posterior marginal distributions of the epidemiological parameters: $\mathcal{R}_e$, $\sigma$, $\psi$ and $\omega$.

#### Sensitivity analysis

As a sensitivity analysis we re-ran the analysis with a slight change to the preprocessing of the time series data: we did not subtract the number of sequences from the case count (this was originally done to guard against double counting these cases), and we computed the credible intervals using the 2.5% and 97.5% quantiles. As can be seen in Tables S3 and S4, the different preprocessing made almost no difference to the estimates of $\mathcal{R}_e$ and only slightly increased the estimates of the rates of observation. This increase is to be expected since under this preprocessing there are additional unsequenced cases present.

| Parameter | Estimate (95% CrI ) |
|---|---|
| $\mathcal{R}_e$ (before 20 April) | $1.62\ (1.48, 1.75)$ |
| $\mathcal{R}_e$ (20 April – 4 May) | $1.01\ (0.83, 1.21)$ |
| $\mathcal{R}_e$ (4 May – 18 May) | $0.80\ (0.57, 1.00)$ |
| $\mathcal{R}_e$ (after 18 May) | $0.04\ (1.63 \times 10^{-3}, 0.19)$ |
| $\sigma$ | $0.11\ (0.10, 0.14)$ |
| $\psi/\sigma$ | $1.76 \times 10^{-3}(9.17 \times 10^{-4}, 2.94 \times 10^{-3})$ |
| $\tilde{\omega}/\sigma$ | $6.99 \times 10^{-3}(3.69 \times 10^{-3}, 0.01)$ |

**Table S4:** Point estimates of epidemiological parameters (median value) and 95% credible intervals under the alternative preprocessing of the time series where we did not subtract the number of sequences from the time series.

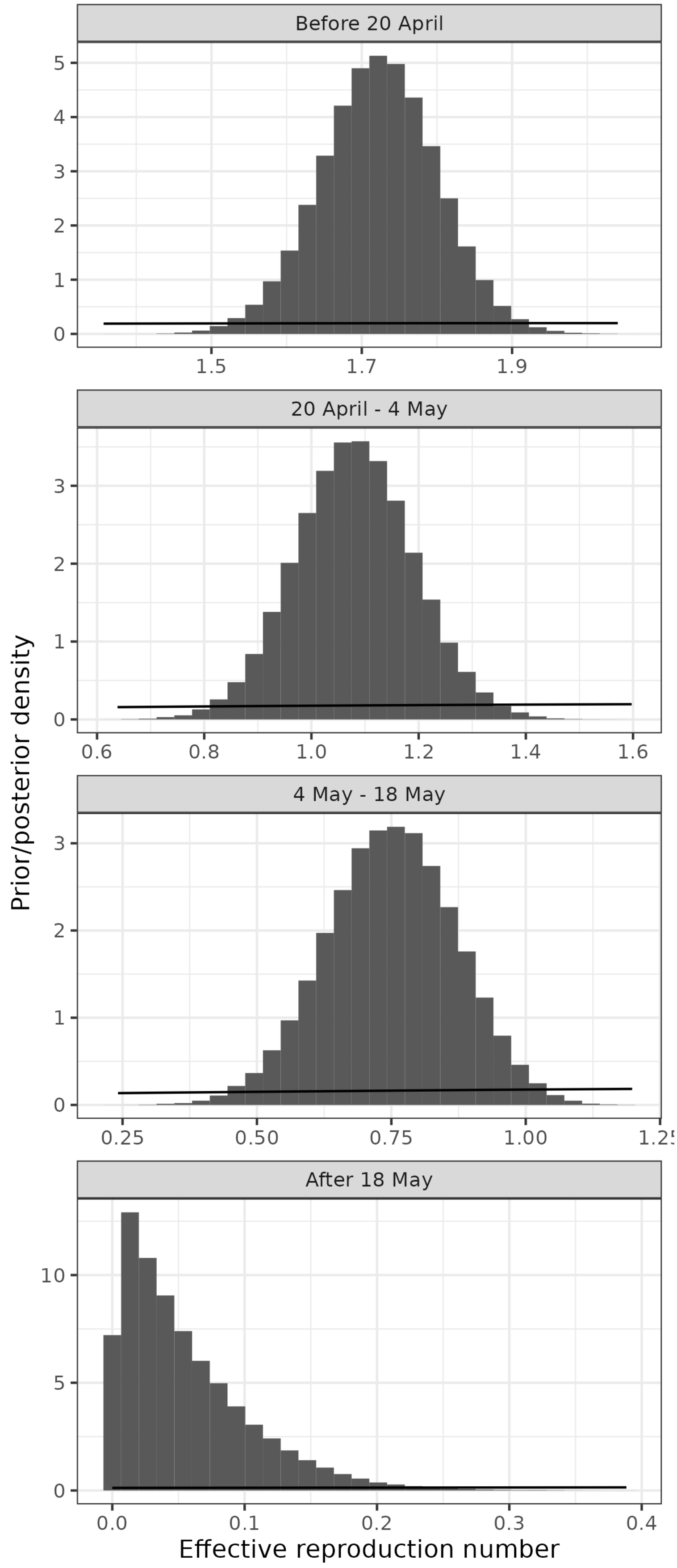


**Figure S4:** The posterior distribution of the effective reproduction number, $\mathcal{R}_{\mathrm{e}}$, in the four time periods. The prior distribution (described in Table S2) is shown with a solid black line.

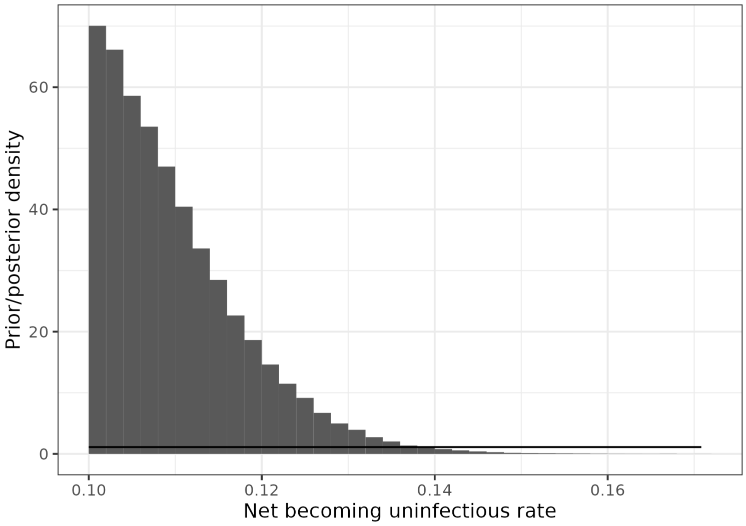


**Figure S5:** The posterior distribution of the net becoming uninfectious rate, $\sigma$. The prior distribution (described in Table S2) is shown with a solid black line.

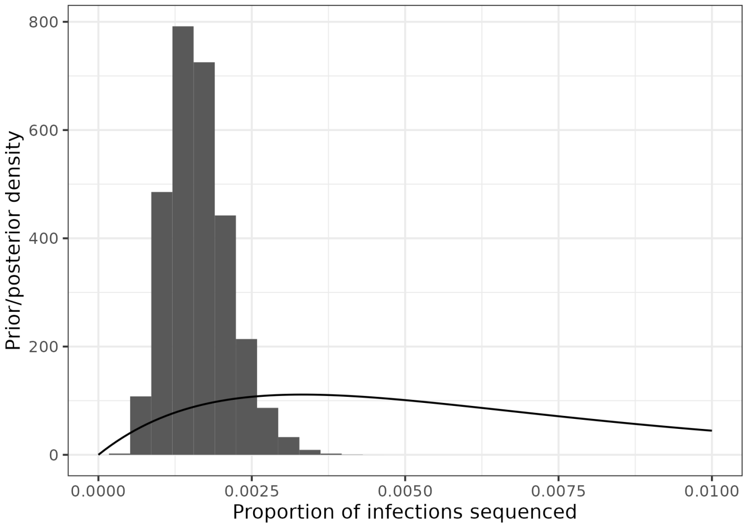


**Figure S6:** The posterior distribution of the proportion of infections that are sequenced, $\psi/\sigma$. The prior distribution (described in Table S2) is shown with a solid black line.

Bouckaert, Remco, Timothy G. Vaughan, Joëlle Barido-Sottani, Sebastián Duchêne, Mathieu Four-

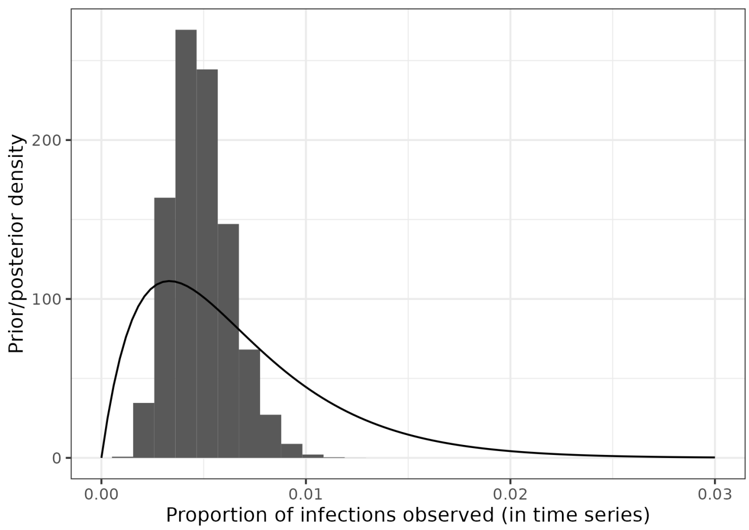


**Figure S7:** The posterior distribution of the proportion of infections that appear in the time series, $\tilde{\omega}/\sigma$. The prior distribution (described in Table S2) is shown with a solid black line.

ment, Alexandra Gavryushkina, Joseph Heled, Graham Jones, Denise Kühnert, Nicola De Maio, Michael Matschiner, Fábio K. Mendes, Nicola F. Müller, Huw A. Ogilvie, Louis du Plessis, Alex Popinga, Andrew Rambaut, David Rasmussen, Igor Siveroni, Marc A. Suchard, Chieh-Hsi Wu, Dong Xie, Chi Zhang, Tanja Stadler, and Alexei J. Drummond (Apr. 2019). "BEAST 2.5: An advanced software platform for Bayesian evolutionary analysis". In: *PLOS Computational Biology* 15.4, pp. 1–28. DOI: 10.1371/journal.pcbi.1006650.

Bouckaert, Remco and Dong Xie (Sept. 2017). *BEAST2-Dev/substmodels: Standard Nucleotide Substitution Models v1.0.1*. Version v1.0.1. DOI: 10.5281/zenodo.995740.

Centers for Disease Control and Prevention (CDC) (2021). *Epidemiology and Prevention of Vaccine-Preventable Diseases*. 14th ed.

Kendall, David G. (1948). "On the Generalized "Birth-and-Death" Process". In: *The Annals of Mathematical Statistics* 19.1, pp. 1–15. DOI: 10.1214/aoms/1177730285.

Li, Lucy M., Nicholas C. Grassly, and Christophe Fraser (July 2017). "Quantifying Transmission Heterogeneity Using Both Pathogen Phylogenies and Incidence Time Series". In: *Molecular Biology and Evolution* 34.11, pp. 2982–2995. DOI: 10.1093/molbev/msx195.

Manceau, Marc, Ankit Gupta, Timothy Vaughan, and Tanja Stadler (2020). "The probability distribution of the ancestral population size conditioned on the reconstructed phylogenetic tree with occurrence data". In: *Journal of Theoretical Biology*, p. 110400. DOI: 10.1016/j.jtbi.2020.110400.

Sekizuka, Tsuyoshi, Kentaro Itokawa, Tsutomu Kageyama, Shinji Saito, Ikuyo Takayama, Hideki Asanuma, Naganori Nao, Rina Tanaka, Masanori Hashino, Takuri Takahashi, Hajime Kamiya, Takuya Yamagishi, Kensaku Kakimoto, Motoi Suzuki, Hideki Hasegawa, Takaji Wakita, and Makoto Kuroda (2020). "Haplotype networks of SARS-CoV-2 infections in the Diamond Princess cruise ship outbreak". In: *Proceedings of the National Academy of Sciences* 117.33, pp. 20198–20201. DOI: 10.1073/pnas.2006824117.

Stadler, Tanja (2010). "Sampling-through-time in birth-death trees". In: *Journal of Theoretical Biology* 267.3, pp. 396–404. DOI: 10.1016/j.jtbi.2010.09.010.

United Nations Statistics Division (2013). *Demographic Yearbook*. Accessed: 2023-10-09. URL: https://unstats.un.org/UNSDWebsite/.

Yakovenko, M L, A P Gmyl, O E Ivanova, T P Eremeeva, A P Ivanov, M A Prostova, O Y Baykova, O V Isaeva, G Y Lipskaya, A K Shakaryan, O M Kew, J M Deshpande, and V I Agol (2014). "The 2010 outbreak of poliomyelitis in Tajikistan: epidemiology and lessons learnt". In: *Eurosurveillance* 19.7. DOI: 10.2807/1560-7917.ES2014.19.7.20706.

Zarebski, Alexander Eugene, Louis du Plessis, Kris Varun Parag, and Oliver George Pybus (Feb. 2022). "A computationally tractable birth-death model that combines phylogenetic and epidemiological data". In: *PLOS Computational Biology* 18.2, pp. 1–22. DOI: 10.1371/journal.pcbi.1009805.
